# The impact of spectral basis set composition on estimated levels of cingulate glutamate and its associations with different personality traits

**DOI:** 10.1101/2023.07.12.23292540

**Authors:** Verena Demler, Elisabeth F. Sterner, Martin Wilson, Claus Zimmer, Franziska Knolle

## Abstract

**Background:** ^1^H-MRS is increasingly used in basic and clinical research to explain brain function and alterations respectively. In psychosis research it is now one of the main tools to investigate imbalances in the glutamatergic system. Interestingly, however, the findings are extremely variable even within patients of similar disease states. One reason may be the variability in analysis strategies, despite suggestions for standardization. Therefore, our study aimed to investigate the extent to which the basis set configuration – which metabolites are included in the basis set used for analysis – would affect the spectral fit and estimated glutamate (Glu) concentrations in the anterior cingulate cortex (ACC), and whether any changes in levels of glutamate would be associated with psychotic-like experiences and autistic traits.

**Methods:** To ensure comparability, we utilized five different exemplar basis sets, used in research, and two different analysis tools, r-based spant applying the ABfit method and *Osprey* using the LCModel.

**Results:** Our findings revealed that the types of metabolites included in the basis set significantly affected the glutamate concentration. We observed that three basis sets led to more consistent results across different concentration types (i.e., absolute Glu in mol/kg, Glx (glutamate+glutamine), Glu/tCr), spectral fit and quality measurements. Interestingly, all three basis sets included phosphocreatine. Importantly, our findings also revealed that glutamate levels were differently associated with both schizotypal and autistic traits depending on basis set configuration and analysis tool, with the same three basis sets showing more consistent results.

**Conclusions:** Our study highlights that scientific results may be significantly altered depending on the choices of metabolites included in the basis set, and with that emphasizes the importance of carefully selecting the configuration of the basis set to ensure accurate and consistent results, when using MR spectroscopy. Overall, our study points out the need for standardized analysis pipelines and reporting.

## 1 Background

Proton Magnetic Resonance Spectroscopy (^1^H-MRS) was developed in the late 1980s and has since then become a powerful tool to measure brain metabolites non-invasively. In many neurological and psychiatric diseases, the brain metabolism is altered, leading to changes in metabolite concentrations across the brain. Therefore, ^1^H-MRS offers a chance for enhancing our understanding of diseases and potentially allowing improvement in developing diagnosis or treatment strategies. By measuring the frequency and intensity of the resonance signals of hydrogen atoms (protons) in certain molecules, ^1^H-MRS can provide information about the types and amounts of chemicals present in the scanned tissue (1). Among many others, glutamate (Glu) and gamma-aminobutyric acid (GABA) – two of the most important neurotransmitters in the brain – can be detected and quantified in the in vivo brain tissue.

^1^H-MRS is an extremely important method for psychiatric research. Studies suggest that an imbalance in different neurotransmitter systems, especially the excitatory glutamatergic and the inhibitory GABAergic systems, contributes to the development of the complex set of symptoms in psychotic disorders and autism spectrum disorder (ASD) (2–6). The glutamate hypothesis of schizophrenia, for example, is based on the finding that psychotic symptoms could be induced by antagonists of a glutamate receptor, specifically the N-methyl-D-aspartate (NMDA) receptor (7,8). Changes in glutamate in the anterior cingulate cortex (ACC) (9) and the bilateral medial prefrontal cortex (10) have been linked to psychotic-like experiences in healthy people as well as to symptoms in both first-episode psychosis (11–13) and chronic psychosis (14). In high-risk individuals, structural changes and symptoms seem to be associated with alterations in ACC glutamate concentrations (11). Similarly, in ASD research, studies found increased glutamate concentrations in the ACC in adolescent autistic males (15) and in children (16).

Due to the relevance of this method for understanding the underlying mechanism of psychiatric disorders, the use of this imaging technique has vastly increased, which is illustrated by an exemplary literature search for ^1^H-MRS and psychotic disorders since the 1980s (Figure 1), and new analysis tools and methods were developed, e.g. FSL-MRS (17), Gannet (18), INSPECTOR (19), Osprey (20), spant (21), and several more. Unfortunately, however, this has resulted in increased variability in the outcomes, which makes it challenging to compare results across studies. Bhogal et al. (22) showed for example, that the metabolite quantification was impacted by the selection of processing parameters and software. But even when using the same software, different processing options had an impact on metabolite quantification. Craven et al. (23) compared seven modelling algorithms for GABA ^1^H-MRS and detected systematic differences for the metabolite estimates between datasets acquired on hardware from different vendors and across algorithms. These and further studies (24–26), assessing the impact of analysis strategies on metabolite quantification, emphasize the importance of creating standards for ^1^H-MRS preprocessing, analysis, and reporting schemes. As a consequence, Lin et al. (27) proposed much needed minimum Reporting Standards for in vivo MRS to enhance the reproducibility of study outcomes and to provide a crucial technical assessment of methodologies and analyses. One important aspect that is often neglected is the impact of the basis set composition on the metabolite levels, and whether potential further analyses using these concentrations show inconsistencies. Here we would like to increase the awareness as many scientists who may conduct mainly clinical research might not know of the importance of selecting metabolites included in the composition of the basis set, and may simply follow departmental routines, which might have been set up for different purposes.

**Figure 1:**
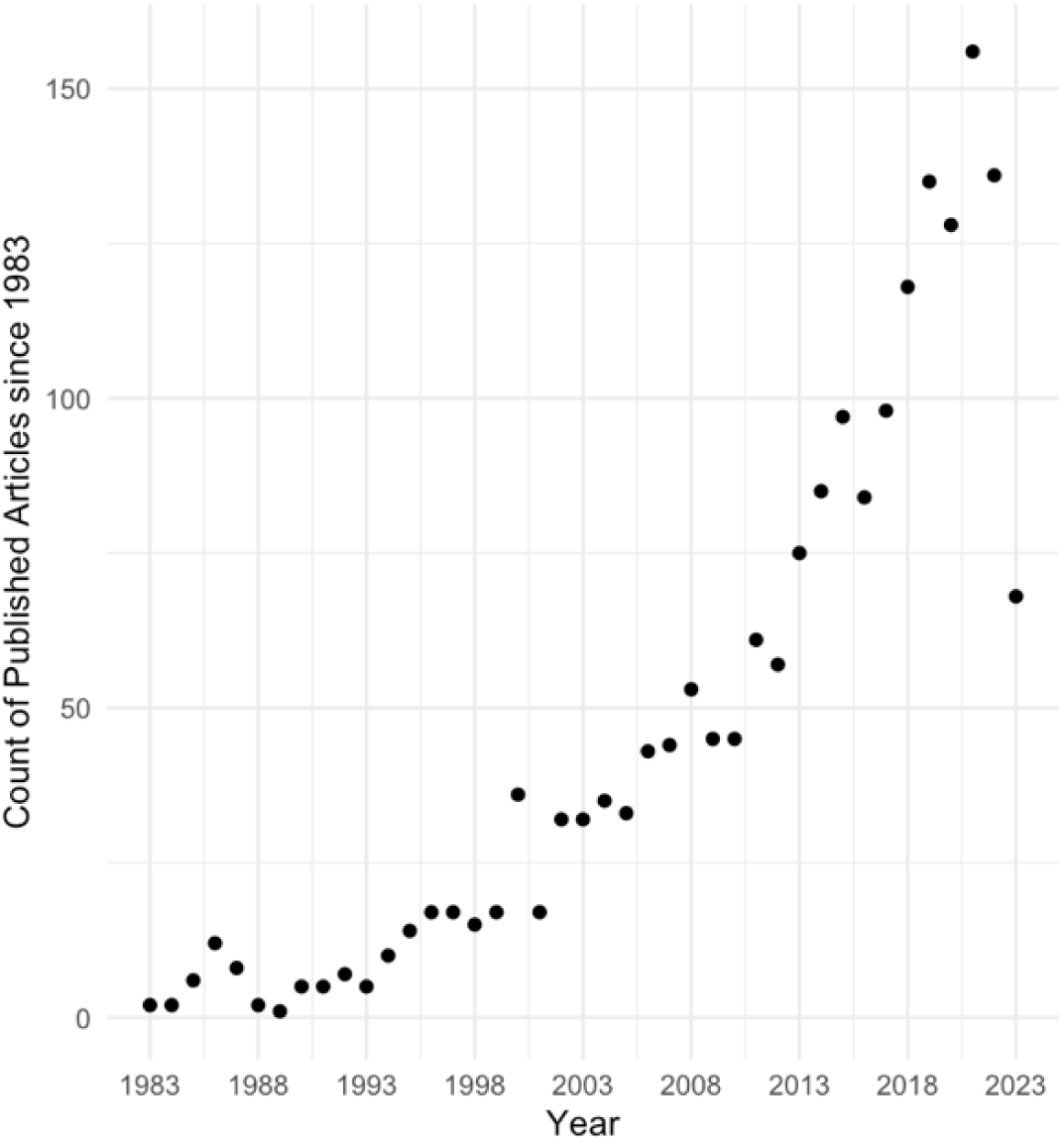
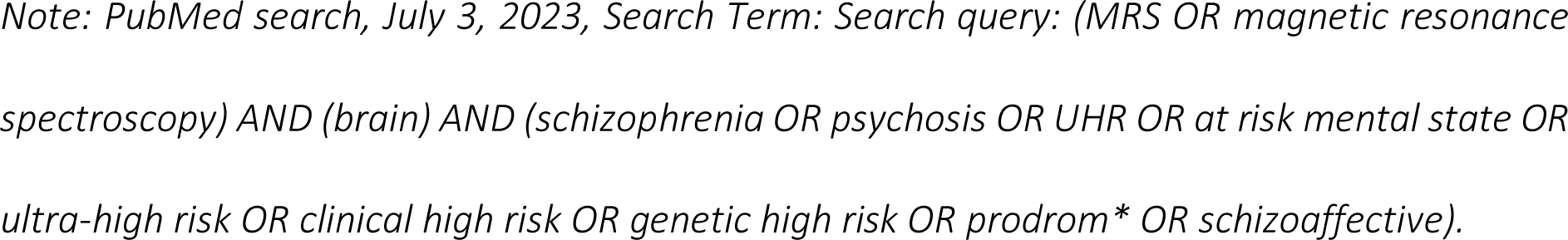
Use of MRS in the research field of psychosis.

As an illustrative example we will concentrate on the investigation of levels of glutamate in patients with psychosis. When investigating levels of glutamate using ^1^H-MRS assessed in psychosis patients, it becomes apparent that there is considerable inconsistency in the results even within the same region. One study reported higher levels (28), whereas others found reductions (29–31) or no significant differences (32,33) in first-episode psychosis or schizophrenic patients in the ACC. The same irregularity can be found for subjects at risk for psychosis, in the ACC higher (4,11,34–36) and lower (10) glutamate levels, or even no significant results (37–39) have been described. Besides the differences between patient groups, even of the same disease stage (e.g., due to medication status), exact voxel placement, and acquisition with different scanner hardware; one problem, adding heterogeneity to the results, could be the choice of the components of a basis set.

A basis set comprises individual metabolite spectra for quantifying acquired signals (i.e., metabolites of interest) and is used to fit the obtained spectrum. Previous studies showed that a basis set used for the analysis of recorded spectroscopy data crucially requires matching acquisition parameters (i.e., pulse sequence, B_0_ field strength, time of echo (TE), spectral bandwidth (BW), and data points) with those of the measured spectrum to enable an accurate fitting result (22,40). Importantly, however, also the selection of metabolites represented in the basis set may affect the fit and concentration of the metabolites of interest. For example, when glutathione (GSH) is excluded from the basis set, those metabolites that have overlapping resonance peaks with GSH, such as GABA, glutamine (Gln), and glutamate (Glu), exhibit notable differences in their concentration (41). Therefore, a potential reason for the diversity in the results regarding levels of glutamate in psychosis could be the choice of basis set components. Indeed, basis sets, if reported, vary grossly between studies and are also describing different results (see Table S1). For example, in a study by Shukla et al. (42) glutamate levels in the ACC covarying for age, were significantly higher in controls compared to patients with schizophrenia, using the following basis set components: alanine (Ala), aspartate (Asp), creatine (Cr), GABA, glucose (Glc), Glu, Gln, GSH, glycine (Gly), glycerophosphocholine (GPC), lactate (Lac), myo-Inositol (mI), N-acetylaspartate (NAA), N-acetylaspartylglutamate (NAAG), phosphocholine (PCh), phosphocreatine (PCr), phosphoryl ethanolamine (PE), scyllo-Inositol (sI) and taurine (Tau). Whereas a study by Rowland et al. (43) on patients with schizophrenia found lower levels of ACC glutamate compared to controls, using these basis set components: Ala, Asp, Cr, GABA, Glc, Gln, Glu, GSH, GPC, Lac, mI, NAA, NAAG, PCh, sI and Tau. Especially for clinical research and the ultimate translation into healthcare, it is crucial that potential causes for variability between studies are detected and that guidelines for standardized ^1^HRS analyses are created. This study, therefore, aims to investigate (1) whether or rather to which extent the choice of metabolites included in the basis set alters spectral fit and/or estimated glutamate concentrations in a voxel; and (2) whether, within and across different basis sets, different representations of glutamate concentrations such as absolute values of glutamate or creatine scales levels of glutamate are differently associated with subclinical traits.

## 2 Materials and Methods

### 2.1 Participants

The study population consisted of 53 healthy subjects (age, 23.6 ± 3.8 years; range, 18–35 years) recruited from the general population in Munich, Germany. All participants (26 women, and 27 men) completed two clinical online questionnaires prior to participating in the study at the Technical University of Munich. To assess their psychotic-like experiences, we applied a German translation (44) of the modified version of the Schizotypal Personality Questionnaire (SPQ) (45) using a 5-point Likert scale version. The German translation of the Autism Spectrum Quotient (AQ) (46) was used to capture the autistic traits. Furthermore, we collected their demographic data and medical history during a brief telephone screening. Additional information regarding the inclusion criteria, demographic data, and symptom scores can be found in the supplementary materials (Table S2). The study was approved by the medical research ethics committee of the Technical University of Munich. All subjects gave written informed consent in accordance with the Declaration of Helsinki.

### 2.2 MR Data acquisition

Structural MRI and 1H-MRS data were acquired using a 32-channel head coil on a 3T Philips Ingenia Elition X MR-Scanner (Philips Healthcare, Best, The Netherlands). We obtained T1- weighted magnetization prepared rapid gradient echo (MPRAGE) images for spectroscopic voxel placement and tissue segmentation (TE, 4ms; Repetition Time (TR), 9ms; Flip angle (α), 8°; shot interval, 3000ms; slice number, 170; matrix size, 240×252 and voxel size, 1×1×1mm³). Single-voxel spectra were collected from a voxel (20×20×20mm^3^) in the ACC. See Figure 2 for voxel placement overlap. Scan parameters for the ECHO volume Point Resolved Spectroscopy Sequence (PRESS) sequence were as follows: TE set to shortest, which resulted in a range of 35.6ms-41.2ms (this is being accounted for in the basis sets); TR, 2000ms; 16 phase cycle steps; acquisition BW, 2 kHz; 1024 data points; flip angle, 90°. To minimize residual water, we used the conventional Philips water suppression technique (excitation) that performs Automatic Water Suppression Optimization (AWSO) pre-scans.

**Figure 2:**
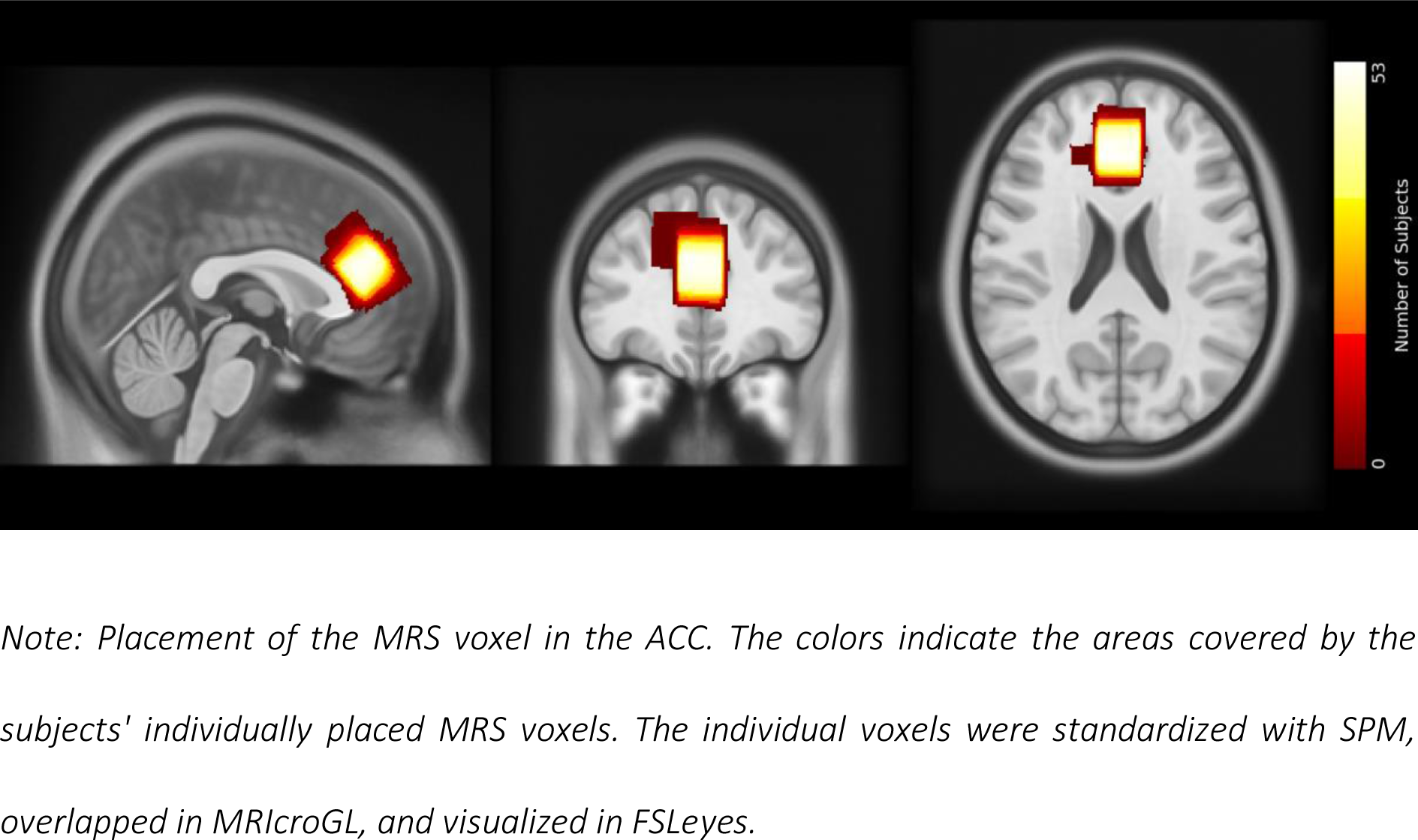
Voxel placement.

### 2.3 1H-MRS Processing

The ^1^H-MRS data were analyzed independently using two different toolboxes: Spectroscopy Analysis Tools (spant) version 2.6.9 (21) (https://martin3141.github.io/spant/index.html) implemented in the open-source R toolbox; and Osprey version 2.4.0, an all-in-one software suite for state-of-the-art processing and quantitative analysis of in-vivo MRS data (20). The scanner’s automatic preprocessing included coil combination, phase-frequency alignment, and averaging of repetitions. In spant, we performed two steps: spectral alignment by referencing the spectrum to the tNAA resonance peak at 2.01 ppm and water removal, which eliminated the residual water signal with a Hankel singular value decomposition (HSVD) filter (47). In Osprey, we excluded eddy-current correction for the automatic preprocessing. Subsequently, Osprey executed the necessary processing steps based on the provided data, including frequency-and-phase alignment, water removal, frequency referencing, and initial phasing. For a more comprehensive overview of the process, see Figure S1.

### 2.4 Basis sets

Due to the varying TE in our ^1^H-MRS data between and within the individual voxels, we utilized MARSS in INSPECTOR version 11-2021 (40) to generate six distinct PRESS basis sets for each TE (36, 37, 38, 39, 40, 41), employing a bandwidth of 2000Hz and 1024pts in each simulation. Based on the literature, we selected five different exemplar basis set compositions for illustration. First, Rowland et al. (48) provides evidence for changes in glutamatergic and GABAergic function in relation to the progression of schizophrenia and the manifestation of cognitive and negative symptoms in individuals with schizophrenia. The study utilized a set of 16 metabolite components: Ala, Asp, Cr, GABA, Glc, Glu, Gln, GSH, GPC, Lac, mI, NAA, NAAG, PCh, sI, and Tau. Second, the standard toolbox for ^1^H-MRS analyses – LCModel – provides a recommendation for the basis set in their manual (49) including 17 metabolites: Ala, Asp, Cr, GABA, Glc, Glu, Gln, GSH, GPC, Lac, mI, NAA, NAAG, PCh, PCr, sI and Tau. Third, Maddock et al. (50) measured glutamate and GABA simultaneously in first-episode psychosis patients and healthy individuals and achieved comparable results for glutamate for the MEGA-PRESS off-resonance to separately-acquired PRESS spectra. They chose 15 metabolites as components of their basis set: Asp, Cr, GABA, Glc, Glu, Gln, GSH, GPC, mI, NAA, NAAG, PCr, PCh, sI, and Tau. Forth, Reid et al. (13) described lower glutamate levels in the ACC in first-episode schizophrenic patients. Their basis set included 19 metabolites: Ala, ascorbate (Asc), Asp, Cr, GABA, Glc, Glu, Gln, GSH, GPC, Lac, mI, NAA, NAAG, PCr, PCh, PE, sI and Tau. Finally, Kozhuharova et al. (10) also simulated 19 basis spectra, which were partially but not fully overlapping with Reid et al. (13), and found lower glutamate levels in high schizotypy individuals in the medial prefrontal cortex, an area often overlapping with the ACC in ^1^H-MRS studies. Their basis set contained: Ala, Asc, Asp, Cr, GABA, Glc, Glu, Gln, glycine (Gly), GSH, GPC, Lac, mI, NAA, NAAG, PE, PCh, sI, and Tau. We added the default macromolecular and lipid components provided by spant or Osprey.

### 2.5 Spectral Fitting

Spant uses an adaptive baseline fitting algorithm (ABfit) (51), which accurately estimates the optimal baseline – hereafter referred to as spant+ABfit. This is important because the smoothness of the baseline is a critical analysis parameter for metabolite estimation. Additionally, spant combines the capabilities of R with a blend of conventional and up-to-date MRS data processing methods, enabling it to perform a fully automated routine MRS analysis (21). We performed the segmentation of the structural T1 image into grey matter (GM), white matter (WM), and cerebrospinal fluid (CSF) using SPM12 (52). For estimation of absolute levels of glutamate (absolute Glu), we utilized the ABfit method to quantify glutamate into a tissue and relaxation-corrected molal glutamate concentration (mol/kg). The correction for tissue fractions was applied using the method described by Gasparovic and colleagues (53).

In Osprey, we used the LCModel (LCM) implementation to fit and quantify our data, hereafter referred to as Osprey+LCM. The LCModel algorithm fits spectra in the frequency domain using a linear combination model (54). The processing adhered to standard parameters, employing a metabolite fit range spanning from 0.5 to 4.0 ppm and a water fit range ranging from 2.0 to 7.4 ppm. A knot spacing of 0.4 ppm was utilized. Osprey calls the SPM12 (52) segmentation function to segment the structural image into tissue probability maps. These are then overlaid with the coregistered voxel masks, created by the Coregistration module, to calculate the fractional tissue volumes for GM, WM, and CSF. For the estimation of absolute levels of glutamate (absolute Glu), Osprey+LCM estimates the tissue and relaxation corrected molal concentration (mol/kg) according to the Gasparovic method (53).

We ran the analysis in both tools for all five basis sets separately. Afterwards, we extracted the scaled estimates for Glu (Glu/tCr), absolute Glu and Glx (Glu+Gln). Note that the basis set of Kozhuharova (10) and Rowland (48) did not include PCr, so the scaled estimates for Glu were calculated to Cr instead of tCr.

### 2.6 Quality assessment

Quality parameters used in this study were the Signal-to-Noise Ratio (SNR), the Full Width at Half Maximum (FWHM) and the Cramer-Rao lower bounds (CRLB). However, we would like to point out that the two analysis tools differently determine some of them. Spant+ABfit calculates the SNR by taking the signal from the maximum point in the fit and subtracting the mean noise value after fitting the data (spant(21): calc_spec_snr). Thus, it is dependent on the baseline intensity. In Osprey+LCM, however, the SNR is calculated by dividing the amplitude of the NAA peak by the standard deviation of the detrended noise in the range of -2 to 0 ppm (20).

Osprey+LCM reports the linewidth as the width of the water peak at half the maximum amplitude calculated as the average of the FWHM of the data and the FWHM of a Lorentzian fit. The threshold was set to the resulting water FWHM > 13 Hz according to recommendations for B0-shimming provided by Juchem et al. (55). In spant+ABfit, the linewidth was given in ppm as tNAA linewidth, full-width half-maximum of a single-Lorentzian fit to the NAA peak (between 1.8 and 2.2 ppm). Based on a consensus paper by Wilson et al. (56), a FWHM greater than 0.1 ppm should be regarded as being of poor quality.

Further exclusion criteria for both analysis methods included either a visual failure of the fitting algorithm or CRLB exceeding 20% for Glu and Glx. Based on these criteria, no subject had to be excluded.

### 2.7 Analysis of group differences between the different basis sets

All statistical analyses were performed using R Statistical Software (version 4.2.2) (57). We tested the different metabolite concentrations across all basis sets for normality using the Shapiro–Wilk method (58) (stats package (57), version 4.2.2) and for homogeneous distribution of their variances by the Levene’s test (59) (cars package (60), version 3.1-1). As the normality assumption and the homogenous distribution of variances were not fulfilled for the metabolite concentrations (Glu/tCr, absolute Glu and Glx) and the spectral quality parameters(Glu CRLB and Glx CRLB), we used the Friedmann test (61) (rstatix package (62), version 0.7.1) to determine group differences across the different basis sets. For post-hoc multiple pairwise-comparison between basis sets, we applied the paired Wilcoxon signed-rank test (63) (rstatix package (62), version 0.7.1). For the group comparison of the SNR, we computed repeated measures ANOVA and multiple pairwise paired t-tests (rstatix package (62), version 0.7.1). P-values were adjusted using the Bonferroni multiple-testing correction method. An adjusted p-value <0.05 was considered significant. The differences were visualized with boxplots with the ggpubr package (64) (version 0.5.0).

### 2.8 Correlation analysis of each metabolite concentration between the basis sets

To investigate the comparability between all our results, we calculated Spearman’s rank correlation coefficients as the normality, linearity, and homoscedasticity assumptions were not met, between the results of the different basis sets and toolboxes for each metabolite estimate (Glu/tCr, absolute Glu, Glx). For these correlation analyses, we used the stats package (57) (version 4.2.2). The correlation heat maps were visualized with the ggcorrplot package (65) (version 0.1.4).

### 2.9 Association between the concentrations and traits

Finally, we explored the relationship between the different metabolite concentrations and clinical scores. First, we factored the SPQ score into positive-like symptoms, negative-like symptoms, and disorganized traits (66). Then, we calculated Spearman’s rank correlation coefficients using the SPQ subscales and autistic traits together with the different metabolite concentrations. The correlation analyses were performed in R using the stats package (57) (version 4.2.2). The visualization of the correlation heat maps was created with the ggcorrplot package (65) (version 0.1.4).

## 3 Results (687)

### 3.1 Spectral quality

All acquired spectra were of good quality with CRLB < 10%, SNR > 60 and for spant FWHM < 0.06 ppm or for Osprey FWHM < 8 Hz. Example fits for the different basis sets are shown in Figure 3. Examining the residuals at the top of every plot, we can see that the fitted models of the basis sets from the LCModel Manual (49), Maddock et al.(50), and Reid et al. (13) had the best fit indicated by lowest fluctuations. Comparing CRLB of Glu and Glx between the basis sets, we found significant overall effects (spant+ABfit: CRLB Glu X^2^ = 40.33, p<0.0001; CRLB Glx: X^2^ = 12.40, p=0.0146; Osprey+LCM: CRLB Glu X^2^ = 17.45, p=0.0016; CRLB Glx: X^2^ = 27.47, p<0.0001) (Table S3). Pairwise post-hoc analyses for CRLB of Glu revealed differences between the basis sets of Rowland et al. (48) and Reid et al. (13) spant+ABfit, and Osprey+LCM and between Maddock (50) and Reid (13) in spant + ABfit (Table S5). Pairwise post-hoc analyses for CRLB of Glx showed significant differences between the basis sets of Maddock et al. (48) and Kozhuharova et al. (10) in spant+ABfit, and Osprey+LCM, and several more in Osprey+LCM (Table S6). Furthermore, we compared SNR between the basis sets (Table S3). In spant + ABfit, but not Osprey, we found differences in SNR among the fitted models (F_1.68, 87.49_ = 31.03, p <0.0001). Lastly, we compared FWHM for spant+ABfit between the basis sets (Table S3), which revealed a significant difference (X^2^ = 150.7, p <0.0001). Post-hoc results are presented in Table S7. FWHM for Osprey+LCM showed identical results across basis sets due to analysis method and was not statistically compared.

**Figure 3:**
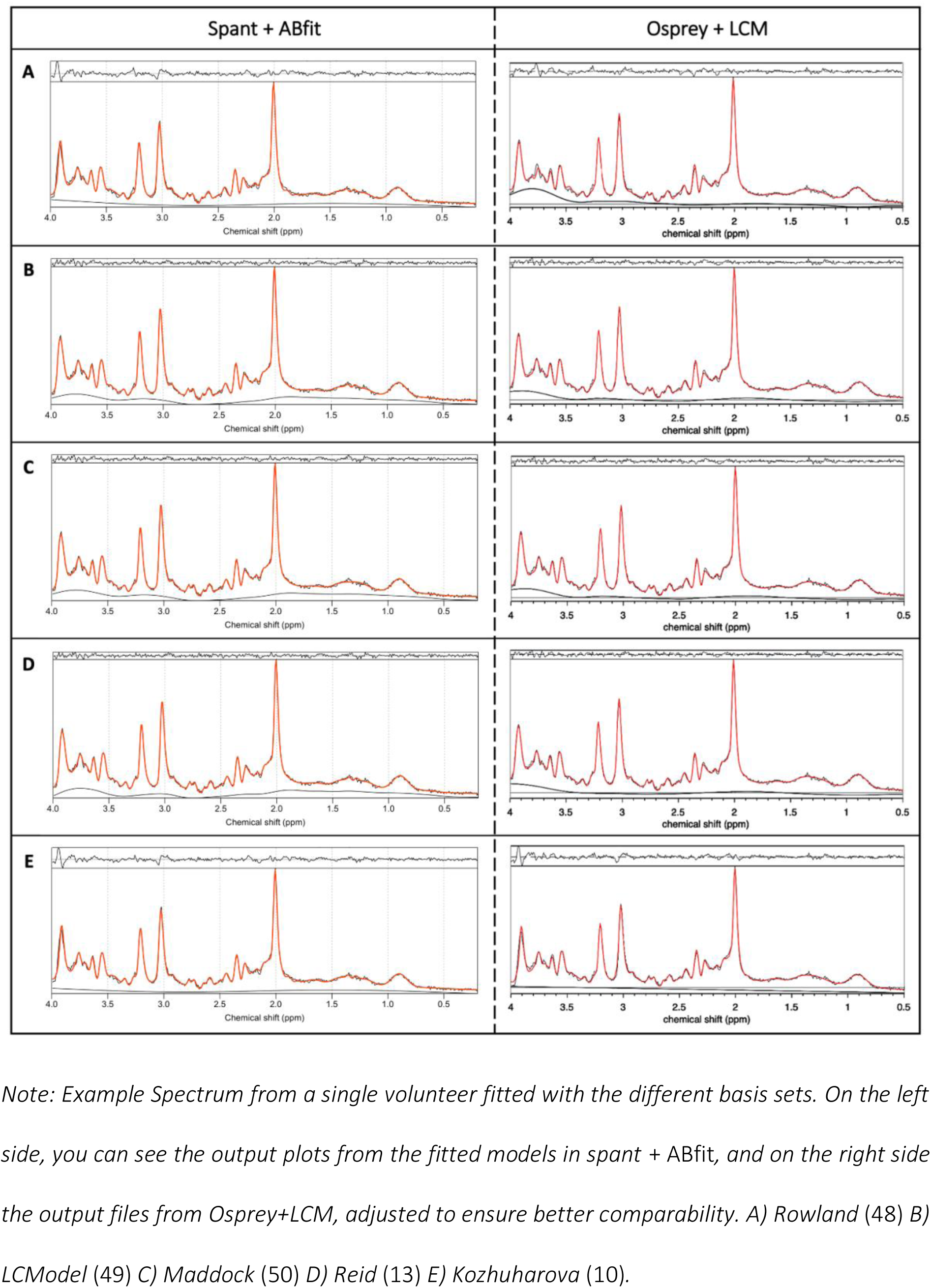
Representative spectrum for each fit.

### 3.2 Group differences between the metabolite concentration estimates

Comparisons between the estimates for Glu/tCr, absolute Glu, and Glx are summarized in Figure 4, results are presented in Table S8-11. The Friedmann test (61) showed significant group differences for t Glu/tCr, absolute Glu, and Glx with always p < 0.0001. Estimates were consistently lower for the analysis in spant+ABfit compared to the ones analyzed with Osprey+LCM. Based on the visualization of the individual data points and their connection over the different basis sets, we found a higher heterogeneity of the metabolite concentration estimates in spant+ABfit compared to Osprey+LCM.

**Figure 4:**
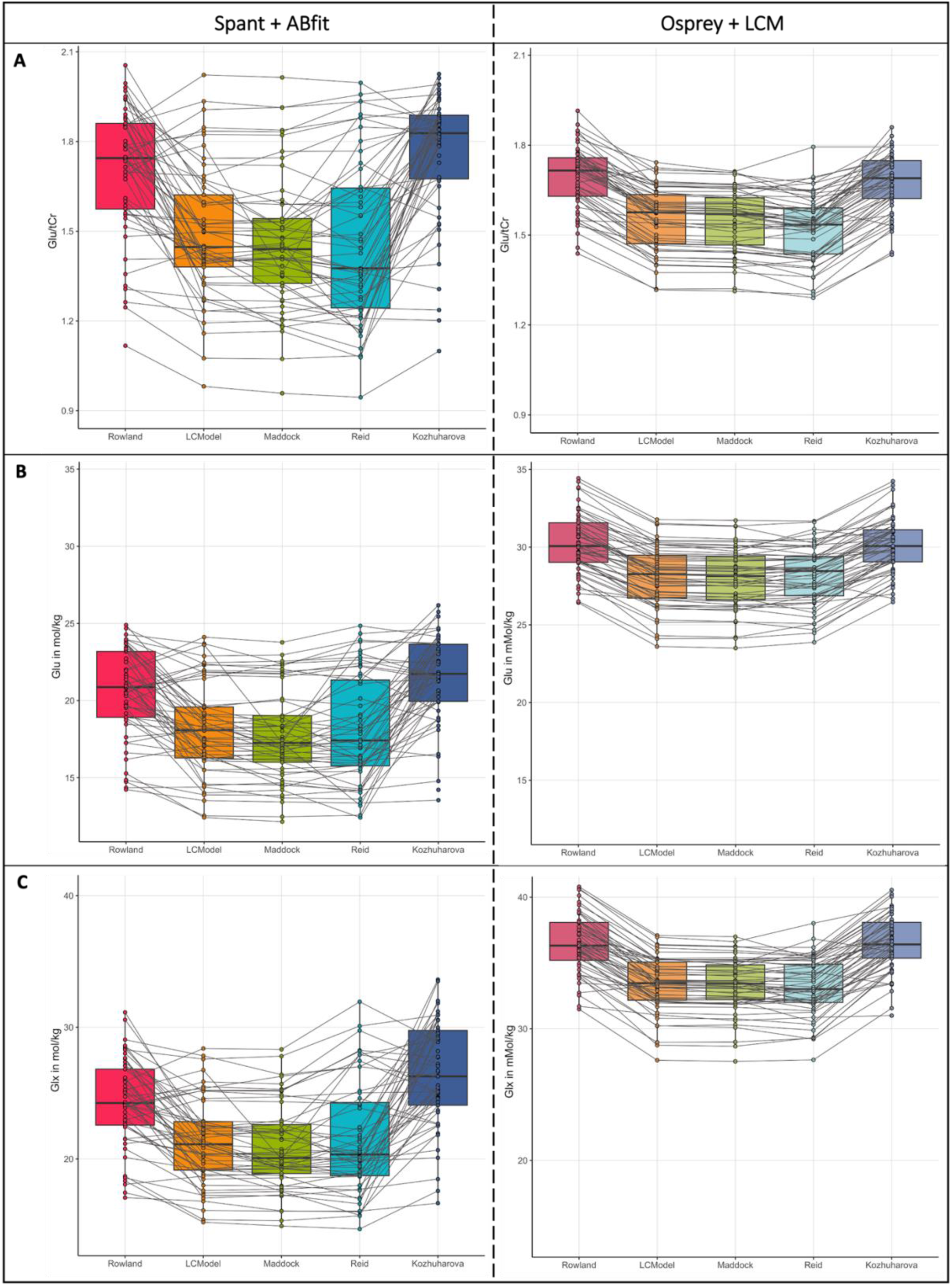
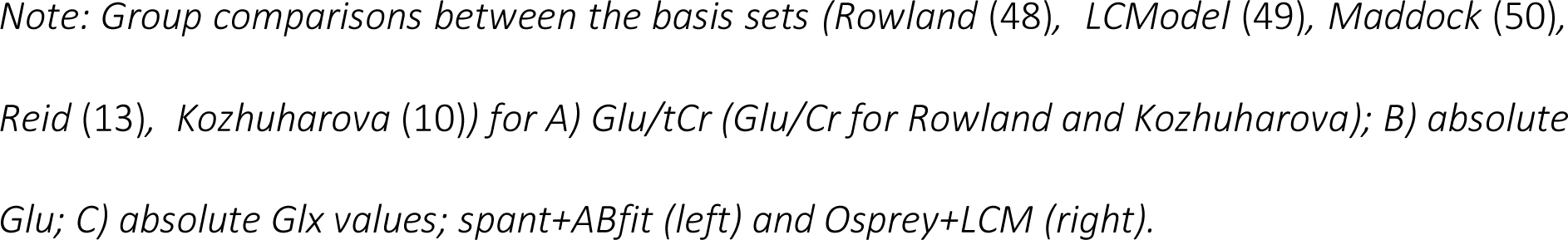
Group comparisons of Glu/tCr, Glu and Glx concentrations within the ACC.

### 3.3 Correlations of each metabolite estimate between the basis sets

Correlations between metabolite (Glu/tCr, absolute Glu, Glx) and basis set per toolbox reveal that the metabolite concentrations between the basis sets had higher correlations using Ospreys LCM integration than spant+ ABfit. For Osprey+LCM, we found strong correlations between all basis sets (r>0.75). The quantification results in spant+ABfit showed a much higher variability. We found weak, moderate, and strong correlations. Overall, the three visually best-fitting basis sets (LCModel Manual (49), Maddock et al. (50), and Reid et al. (13); Figure 3 B, C, and D) showed the best results with spearman correlation coefficients between 0.93-1 for Osprey+LCM and 0.88-0.94 for spant+ABfit (Figure 5). Another notable aspect is, that the two basis sets Rowland (48) and Kozhuharova (10), which both do not include PCr in their basis set composition, showed a strong correlation within each analysis tool, Osprey+LCM and spant+ABfit. Between the toolboxes, the correlations were weak to moderate. Correlation strength was classified according to Akoglu (67).

**Figure 5:**
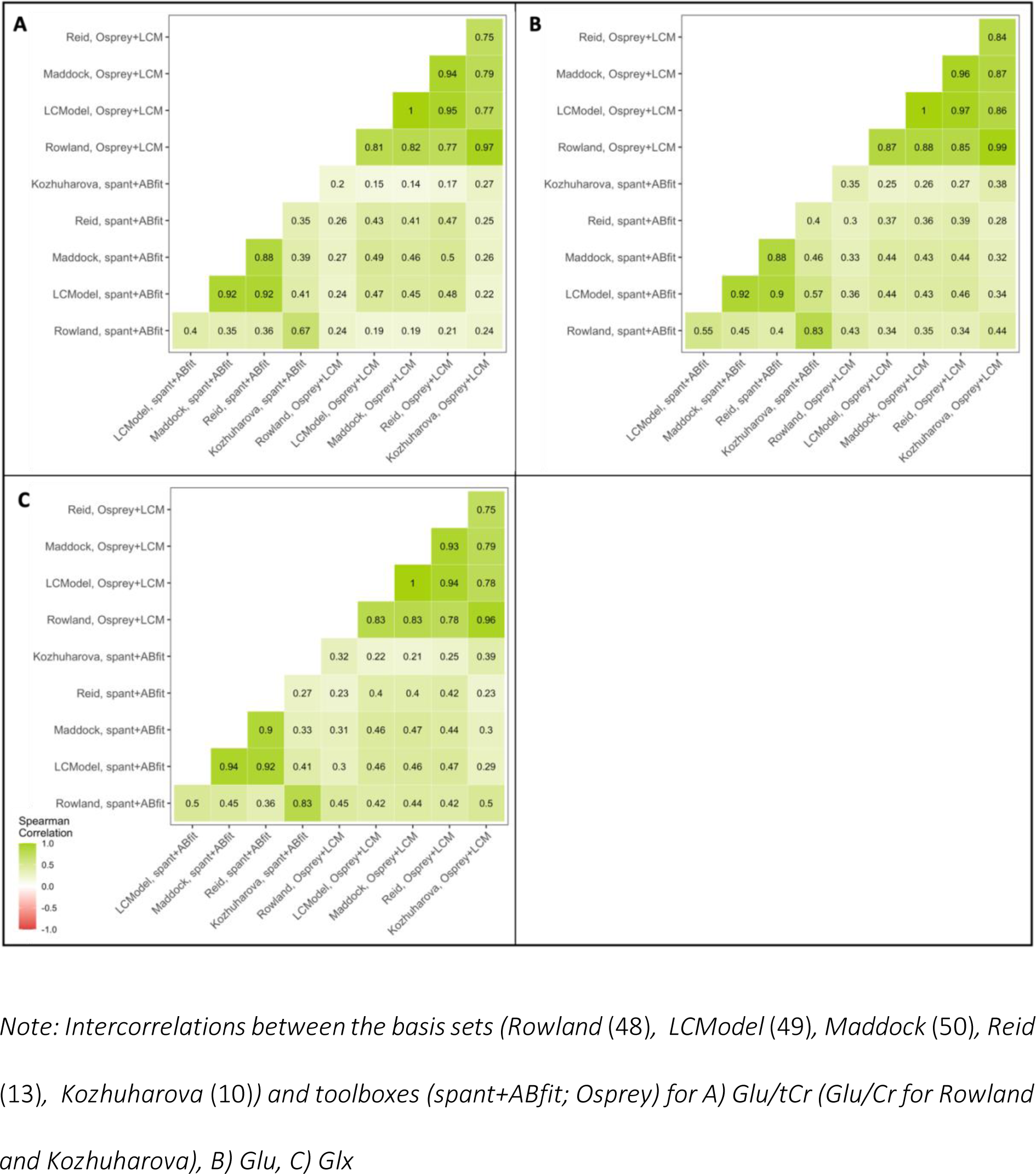
Basis set and tool box intercorrelations.

### 3.4 Association between the concentrations and traits

Finally, we analyzed Spearman’s rank correlations for clinical scores with the extracted concentration scores. Overall, the results in Osprey+LCM displayed a more homogenous pattern regarding the tendency of their non-significant correlations with a maximum difference of 0.18 between the coefficient scores for the correlation of Glx and disorganized traits, whereas in spant+ABfit the correlations coefficients showed greater variability ranging from positive to close to negative values, with a maximal difference of 0.42 for the same correlation. These results also indicated that the correlation coefficients differed between the two toolboxes. However, the difference between the toolboxes was smaller than the difference across the different basis sets, and reached a maximum of 0.29 again for the correlation of Glx and disorganized traits. The correlations are shown in Figure 6.

**Figure 6:**
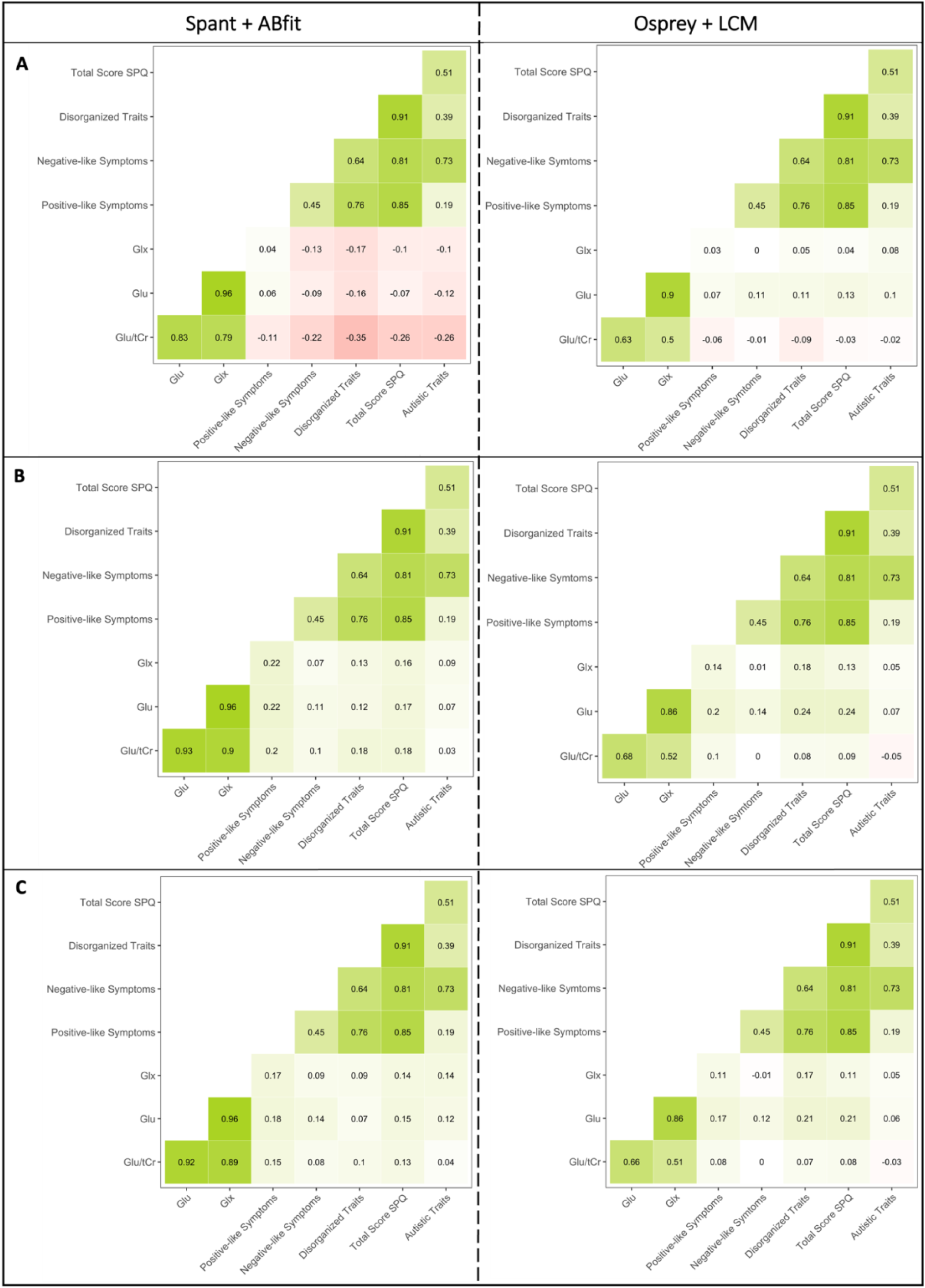
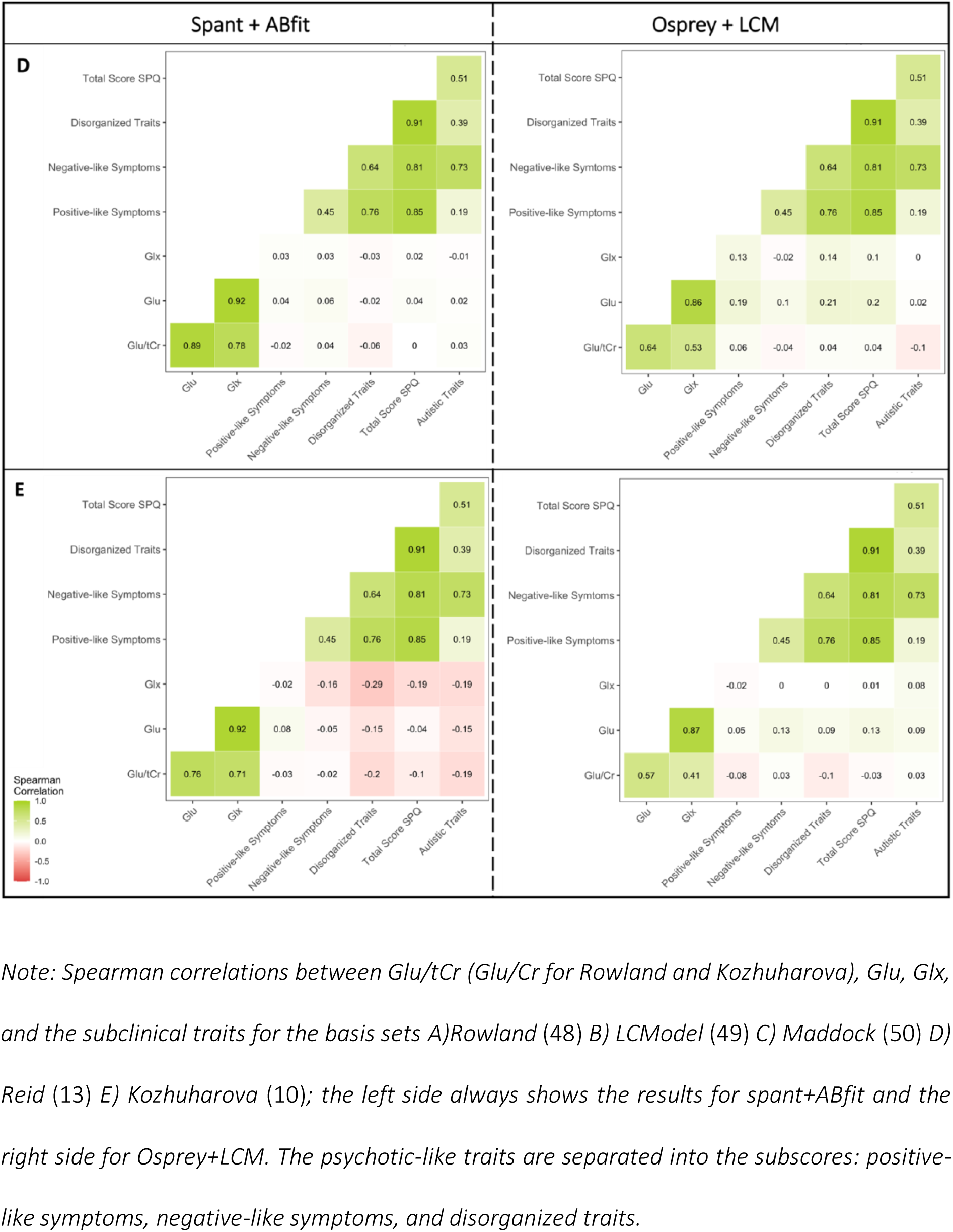
Correlations between the metabolite concentrations and clinical scores.

## 4 Discussion

In this study, we investigated whether the choice of metabolites included in the basis set for a ^1^H-MRS analysis alters spectral fit and/or estimated glutamate concentrations in a voxel placed in the ACC. Importantly, we examined whether potential changes in glutamate concentrations were differently associated with schizotypal and autistic traits. For comparability, we contrasted the effects using five basis sets used in psychosis research and two different analysis tools (i.e., spant+ABfit, Osprey+LCM). We found that (1) glutamate concentrations differed significantly depending on the metabolites included in the basis set with those including phosphocreatine (PCr) showing more consistent results; (2) differences in concentration between the basis sets were similar across the different types of concentration estimates (i.e., absolute Glu, Glx, Glu/tCr); (3) fit and quality measures revealed differences, again with those including phosphocreatine showing better results; and finally (4) concentrations estimated based on different basis sets led to varying symptom correlations, with differences between basis sets being larger than between toolbox, although, Osprey+LCM showed greater consistency in the results compared to spant+ABfit.

Our results indicate that those basis sets containing PCr (i.e., Reid (13), Maddock (50), LCModel Manual (49)) showed better spectral fit and more consistent levels of glutamate. This effect was similar across the different types of concentrations (i.e., absolute Glu, Glx, Glu/tCr). This may be due to the robustness and well-defined chemical shift of the PCr within the MRS spectra of a voxel, which may be caused by its a relatively high and stable concentrations across the brain (68,69). Therefore, basis sets which did not include PCr may produce more variability in the data due to a worse fit. Interestingly, the inclusion of PCr seems to override differences regarding to the number of metabolites included between the basis sets, as Maddock et al (50) include four spectra less compared to Reid et al (13). This result was consistent between both toolboxes, spant+ABfit and Osprey+LCM. To test this, we added the metabolite PCr to the basis sets of Rowland (48) and Kozhuharova (10) and re-ran the analyses. The new Rowland basis set with PCr was now identical to the basis set of the LCM Manual. Interestingly, after adding PCr, both basis sets produced comparable results to those basis sets containing PCr from the beginning (i.e., Reid (13), Maddock (50), LCModel Manual (49)), see Figure S2 and S3. Here, our results seem to identify PCr as a crucial metabolite for the fitting of different glutamate concentrations (i.e., absolute Glu, Glx, Glu/tCr).

This result was confirmed by the finding that those three basis sets containing PCr also showed the strongest intercorrelations (r=0.88-1) across all different concentration types within the same toolbox. Across the toolboxes, correlations were weaker. However, the three basis sets containing PCr showed the strongest (r=0.39-0.47) and most consistent correlations across the three glutamate concentrations. Furthermore, those three basis sets also showed significantly less variation compared to the other two basis sets.

When comparing SNR between the basis sets, we found differences in spant+ABfit, but not in Osprey+LCM, where values are identical across the fitted models. This may be explained by the different methods used for SNR calculation. These processing differences may account for the fact that differences were detected in spant+ABfit but not Osprey+LCM.

Considering the results of this study and the inconsistencies reported in the literature regarding, for example, differences in glutamate concentrations in psychosis patients (4,70,71), one pressing question is whether the latter variability is further increased or partially caused by the differences in analysis strategies and parameters, for example the choice of metabolites included in the basis set but also the analysis software or toolbox itself. While there is further variation due to voxel size and placement, analysis software, analysis parameters and field strength (29–31), the configuration of the basis set is easily controllable, and would potentially reduce the variability in the results across studies, for example in psychosis research (29–31). For different measures of glutamate (i.e., absolute Glu, Glx, Glu/tCr), this study indicates that the inclusion of PCr is crucial. However, more research is needed to confirm these results especially using clinical populations. This emphasizes once more that guidelines for ^1^H-MRS analysis (27,55,56,72), especially the configuration for basis sets to estimate concentrations of much studied metabolites, such as glutamate, are essential in order to produce reliable research that allows translational and clinical implications. The findings discussed here are highly important as treatment options are being discussed (29,73) on the basis of findings that may be impacted by analysis choices.

This study has several limitations. First, we analyzed only one voxel in the ACC, and chose five different, exemplar basis sets and we used two different analysis software tools. Although, those choices are not comprehensive, they provide some important insights and they were motivated based on published research, and demonstrate possible analysis strategies. Future research should replicate these analyses in different brain areas, and potentially using additional basis sets and analysis tools. However, the most insightful strategies would be to run a phantom study, which assess the accuracy and stability of levels of glutamate with different basis sets (74). Second, we used personality traits in a healthy general population sample.

Therefore, the distribution of a clinical expression of symptom scores is skewed, and possible effects may be less prominent compared to potential effects in clinical groups. Interestingly, however, even with trait makers that are relatively low our results show differences of the magnitude of a weak to moderate correlation, indicating the impact of analysis choices. Third, our sample was moderate in size with 53 subjects. With larger cohorts, results may stabilize. Nevertheless, one should consider that many patient studies have smaller sample sizes with cohorts of 20-30 subjects. Therefore, it is extremely valuable to show the impact of analysis choices on smaller cohorts. Fourth, we did not calculate test-retest reliability for evaluating individual differences and establishing a certain robustness of our results as we did not conduct a follow-up measurement. This and an experimental quantification of the glutamate would be needed to objectively state the best basis set composition. Fifth, as the focus of this study was to increase the awareness of the matter, we are unable to provide specific recommendations regarding the aspects of preprocessing and analysis methods. However, future studies specifically designed to assess the best fitting basis set composition should provide those, that ought to be part of the minimum Reporting Standards for in vivo Magnetic Resonance Spectroscopy.

In conclusion, our study demonstrates the importance of using consistent basis set compositions for accurate spectral fitting but especially for comparable generation of metabolite estimates. This requires a standardized analysis approach. Potential consequences of the variability in the analysis techniques and strategies used in current research become apparent in the differences of symptoms correlations depending on analysis choices. Therefore, this study once more emphasizes the need for standardized analysis and reporting guidelines in spectroscopy but also imaging research in general.

## 5 Declarations

### Ethics approval and consent to participate

The medical research ethics committee of the Technical University of Munich gave ethical approval for this work. All subjects gave written informed consent in accordance with the Declaration of Helsinki.

### Consent for publication

Not applicable

### Availability of data and materials

The datasets generated and/or analysed during the current study are available in the OSF repository, [https://osf.io/zjtd8/?view_only=9763e1e752e7409892730ecafed71bf7].

### Authors’ contributions

VFD, FK, CZ conceptualization; VFD, FK, EFS data collection; VFD, FK formal data analysis; MW advise on formal analysis; FK, VFD writing for first draft; VFD, FK, MW, EFS, CZ editing and reviewing of manuscript.

## Acknowledgements

We would like to thank all participants for their time and engagement.

## 6 List of abbreviations

^1^H-MRS: Proton Magnetic Resonance Spectroscopy
ABfit: Adaptive baseline fitting algorithm
ACC: Anterior cingulate cortex
AQ: Autism Spectrum Quotient
ASD: Autism spectrum disorder
Asp: Aspartate
AWSO: Automatic Water Suppression Optimization
BW: Spectral bandwidth
Cr: Creatine
CRLB: Cramer-Rao lower bounds
CSF: Cerebrospinal fluid
FWHM: Full Width at Half Maximum
GABA: Gamma-aminobutyric acid
Glc: Glucose
Gln: Glutamine
Glu: Glutamate
Gly: Glycine
GM: Grey matter
GPC: Glycerophosphocholine
GSH: Glutathione
HSVD: Hankel singular value decomposition
Lac: Lactate
LCM: LCModel
MI: Myo-Inositol
MPRAGE: Magnetization prepared rapid gradient echo
MRS: Magnetic Resonance Spectroscopy
NAA: N-acetylaspartate
NAAG: N-acetylaspartylglutamate
NMDA: N-methyl-D-aspartate
PCh: Phosphocholine
PCr: Phosphocreatine
PE: Phosphoryl ethanolamine
PRESS: Point Resolved Spectroscopy Sequence
SI: Scyllo-Inositol
SNR: Signal-to-Noise Ratio
spant: Spectroscopy Analysis Tools
SPQ: Schizotypal Personality Questionnaire
Tau: Taurine
TE: Time of echo
TR: Repetition Time
WM: White matter

## References

1. Jansen JFA, Backes WH, Nicolay K, Kooi ME. ^1^ H MR Spectroscopy of the Brain: Absolute Quantification of Metabolites. Radiology. 2006 Aug;240(2):318–32.

2. Tebartz van Elst L, Maier S, Fangmeier T, Endres D, Mueller GT, Nickel K, et al. Disturbed cingulate glutamate metabolism in adults with high-functioning autism spectrum disorder: evidence in support of the excitatory/inhibitory imbalance hypothesis. Mol Psychiatry. 2014 Dec;19(12):1314–25.

3. Kolodny T, Schallmo MP, Gerdts J, Edden RAE, Bernier RA, Murray SO. Concentrations of Cortical GABA and Glutamate in Young Adults With Autism Spectrum Disorder. Autism Res Off J Int Soc Autism Res. 2020 Jul;13(7):1111–29.

4. Wenneberg C, Glenthøj BY, Hjorthøj C, Buchardt Zingenberg FJ, Glenthøj LB, Rostrup E, et al. Cerebral glutamate and GABA levels in high-risk of psychosis states: A focused review and meta-analysis of 1H-MRS studies. Schizophr Res. 2020 Jan;215:38–48.

5. Ford TC, Nibbs R, Crewther DP. Glutamate/GABA+ ratio is associated with the psychosocial domain of autistic and schizotypal traits. PLOS ONE. 2017 Jul 31;12(7):e0181961.

6. Sydnor VJ, Roalf DR. A meta-analysis of ultra-high field glutamate, glutamine, GABA and glutathione 1HMRS in psychosis: Implications for studies of psychosis risk. Schizophr Res. 2020 Dec 1;226:61–9.

7. McCutcheon RA, Krystal JH, Howes OD. Dopamine and glutamate in schizophrenia: biology, symptoms and treatment. World Psychiatry. 2020;19(1):15–33.

8. Uno Y, Coyle JT. Glutamate hypothesis in schizophrenia. Psychiatry Clin Neurosci. 2019;73(5):204–15.

9. Modinos G, McLaughlin A, Egerton A, McMullen K, Kumari V, Barker GJ, et al. Corticolimbic hyper-response to emotion and glutamatergic function in people with high schizotypy: a multimodal fMRI-MRS study. Transl Psychiatry. 2017 Apr 4;7(4):e1083–e1083.

10. Kozhuharova P, Diaconescu AO, Allen P. Reduced cortical GABA and glutamate in high schizotypy. Psychopharmacology (Berl). 2021 Sep;238(9):2459–70.

11. Merritt K, Egerton A, Kempton MJ, Taylor MJ, McGuire PK. Nature of Glutamate Alterations in Schizophrenia: A Meta-analysis of Proton Magnetic Resonance Spectroscopy Studies. JAMA Psychiatry. 2016 Jul 1;73(7):665–74.

12. Sigvard AK, Bojesen KB, Ambrosen KS, Nielsen MØ, Gjedde A, Tangmose K, et al. Dopamine Synthesis Capacity and GABA and Glutamate Levels Separate Antipsychotic-Naïve Patients With First-Episode Psychosis From Healthy Control Subjects in a Multimodal Prediction Model. Biol Psychiatry Glob Open Sci. 2022 May;S2667174322000659.

13. Reid MA, Salibi N, White DM, Gawne TJ, Denney TS, Lahti AC. 7T Proton Magnetic Resonance Spectroscopy of the Anterior Cingulate Cortex in First-Episode Schizophrenia. Schizophr Bull. 2019 Jan 1;45(1):180–9.

14. Marsman A, van den Heuvel MP, Klomp DWJ, Kahn RS, Luijten PR, Hulshoff Pol HE. Glutamate in schizophrenia: a focused review and meta-analysis of ^1^H-MRS studies. Schizophr Bull. 2013 Jan;39(1):120–9.

15. Joshi G, Biederman J, Wozniak J, Goldin RL, Crowley D, Furtak S, et al. Magnetic resonance spectroscopy study of the glutamatergic system in adolescent males with high-functioning autistic disorder: a pilot study at 4T. Eur Arch Psychiatry Clin Neurosci. 2013 Aug;263(5):379–84.

16. Naaijen J, Lythgoe DJ, Amiri H, Buitelaar JK, Glennon JC. Fronto-striatal glutamatergic compounds in compulsive and impulsive syndromes: A review of magnetic resonance spectroscopy studies. Neurosci Biobehav Rev. 2015 May 1;52:74–88.

17. Clarke WT, Stagg CJ, Jbabdi S. FSL-MRS: An end-to-end spectroscopy analysis package. Magn Reson Med. 2021 Jun;85(6):2950–64.

18. Edden RAE, Puts NAJ, Harris AD, Barker PB, Evans CJ. Gannet: A batch-processing tool for the quantitative analysis of gamma-aminobutyric acid–edited MR spectroscopy spectra. J Magn Reson Imaging. 2014;40(6):1445–52.

19. Gajdošík M, Landheer K, Swanberg KM, Juchem C. INSPECTOR: free software for magnetic resonance spectroscopy data inspection, processing, simulation and analysis. Sci Rep. 2021 Jan 22;11(1):2094.

20. Oeltzschner G, Zöllner HJ, Hui SCN, Mikkelsen M, Saleh MG, Tapper S, et al. Osprey: Open-source processing, reconstruction & estimation of magnetic resonance spectroscopy data. J Neurosci Methods. 2020 Sep 1;343:108827.

21. Wilson M. spant: An R package for magnetic resonance spectroscopy analysis. J Open Source Softw. 2021 Nov 2;6(67):3646.

22. Bhogal AA, Schür RR, Houtepen LC, van de Bank B, Boer VO, Marsman A, et al. 1H–MRS processing parameters affect metabolite quantification: The urgent need for uniform and transparent standardization. NMR Biomed. 2017;30(11):e3804.

23. Craven AR, Bhattacharyya PK, Clarke WT, Dydak U, Edden RAE, Ersland L, et al. Comparison of seven modelling algorithms for γ-aminobutyric acid–edited proton magnetic resonance spectroscopy. NMR Biomed [Internet]. 2022 Jul [cited 2022 Oct 10];35(7). Available from: https://onlinelibrary.wiley.com/doi/10.1002/nbm.4702

24. Kanowski M, Kaufmann J, Braun J, Bernarding J, Tempelmann C. Quantitation of simulated short echo time 1H human brain spectra by LCModel and AMARES. Magn Reson Med. 2004;51(5):904–12.

25. Zöllner HJ, Považan M, Hui SCN, Tapper S, Edden RAE, Oeltzschner G. Comparison of different linear-combination modeling algorithms for short-TE proton spectra. NMR Biomed. 2021;34(4):e4482.

26. Mikkelsen M, Bhattacharyya P, Mandal P, Shukla D, Wang A, Wilson M, et al. Analyzing Big GABA: Comparison of Five Software Packages for GABA-Edited MRS. 2019.

27. Lin A, Andronesi O, Bogner W, Choi IY, Coello E, Cudalbu C, et al. Minimum Reporting Standards for in vivo Magnetic Resonance Spectroscopy (MRSinMRS): Experts’ consensus recommendations. NMR Biomed. 2021;34(5):e4484.

28. Théberge J, Williamson KE, Aoyama N, Drost DJ, Manchanda R, Malla AK, et al. Longitudinal grey-matter and glutamatergic losses in first-episode schizophrenia. Br J Psychiatry. 2007 Oct;191(4):325–34.

29. Godlewska BR, Minichino A, Emir U, Angelescu I, Lennox B, Micunovic M, et al. Brain glutamate concentration in men with early psychosis: a magnetic resonance spectroscopy case–control study at 7 T. Transl Psychiatry. 2021 Jun;11(1):367.

30. Wang AM, Pradhan S, Coughlin JM, Trivedi A, DuBois SL, Crawford JL, et al. Assessing Brain Metabolism With 7-T Proton Magnetic Resonance Spectroscopy in Patients With First-Episode Psychosis. JAMA Psychiatry. 2019 Mar 1;76(3):314–23.

31. Jeon P, Limongi R, Ford SD, Mackinley M, Dempster K, Théberge J, et al. Progressive Changes in Glutamate Concentration in Early Stages of Schizophrenia: A Longitudinal 7-Tesla MRS Study. Schizophr Bull Open. 2021 Jan 1;2(1):sgaa072.

32. Limongi R, Jeon P, Théberge J, Palaniyappan L. Counteracting Effects of Glutathione on the Glutamate-Driven Excitation/Inhibition Imbalance in First-Episode Schizophrenia: A 7T MRS and Dynamic Causal Modeling Study. Antioxid Basel Switz. 2021 Jan 8;10(1):75.

33. Posporelis S, Coughlin JM, Marsman A, Pradhan S, Tanaka T, Wang H, et al. Decoupling of brain temperature and glutamate in recent-onset of schizophrenia: a 7 Tesla 1H-MRS study. Biol Psychiatry Cogn Neurosci Neuroimaging. 2018 Mar;3(3):248–54.

34. Demro C, Rowland L, Wijtenburg SA, Waltz J, Gold J, Kline E, et al. Glutamatergic metabolites among adolescents at risk for psychosis. Psychiatry Res. 2017 Nov;257:179–85.

35. de la Fuente-Sandoval C, Reyes-Madrigal F, Mao X, León-Ortiz P, Rodríguez-Mayoral O, Solís-Vivanco R, et al. Cortico-Striatal GABAergic and Glutamatergic Dysregulations in Subjects at Ultra-High Risk for Psychosis Investigated with Proton Magnetic Resonance Spectroscopy. Int J Neuropsychopharmacol. 2015 Sep 12;19(3):pyv105.

36. Demler VF, Sterner EF, Wilson M, Zimmer C, Knolle F. Association between increased anterior cingulate glutamate and psychotic-like symptoms, but not autistic traits [Internet]. medRxiv; 2023 [cited 2023 Mar 6]. p. 2023.02.01.23285183. Available from: https://www.medrxiv.org/content/10.1101/2023.02.01.23285183v1

37. Modinos G, Egerton A, McLaughlin A, McMullen K, Kumari V, Lythgoe DJ, et al. Neuroanatomical changes in people with high schizotypy: Relationship to glutamate levels. Psychol Med. 2018 Aug;48(11):1880–9.

38. Egerton A, Stone JM, Chaddock CA, Barker GJ, Bonoldi I, Howard RM, et al. Relationship Between Brain Glutamate Levels and Clinical Outcome in Individuals at Ultra High Risk of Psychosis. Neuropsychopharmacology. 2014 Nov;39(12):2891–9.

39. Yoo SY, Yeon S, Choi CH, Kang DH, Lee JM, Shin NY, et al. Proton magnetic resonance spectroscopy in subjects with high genetic risk of schizophrenia: Investigation of anterior cingulate, dorsolateral prefrontal cortex and thalamus. Schizophr Res. 2009 Jun 1;111(1):86– 93.

40. Landheer K, Swanberg KM, Juchem C. Magnetic resonance Spectrum simulator (MARSS), a novel software package for fast and computationally efficient basis set simulation. NMR Biomed. 2021;34(5):e4129.

41. Hofmann L, Slotboom J, Jung B, Maloca P, Boesch C, Kreis R. Quantitative 1H-magnetic resonance spectroscopy of human brain: Influence of composition and parameterization of the basis set in linear combination model-fitting. Magn Reson Med. 2002;48(3):440–53.

42. Shukla DK, Wijtenburg SA, Chen H, Chiappelli JJ, Kochunov P, Hong LE, et al. Anterior Cingulate Glutamate and GABA Associations on Functional Connectivity in Schizophrenia. Schizophr Bull. 2019 Apr 25;45(3):647–58.

43. Rowland L, Krause BW, Wijtenburg SA, Mcmahon R, Chiappelli J, Nugent K, et al. Medial frontal GABA is lower in older schizophrenia: A MEGA-PRESS with macromolecule suppression study. Mol Psychiatry. 2015 Mar 31;21.

44. Klein C, Andresen B, Jahn T. Erfassung der schizotypen Persönlichkeit nach DSM-III-R: Psychometrische Eigenschaften einer autorisierten deutschsprachigen Übersetzung des ‘Schizotypal Personality Questionnaire’ (SPQ) von Raine. [Psychometric assessment of the schizotypal personality according to DSM-III-R criteria: Psychometric properties of an authorized German translation of Raine’s ‘Schizotypal Personality Questionnaire’ (SPQ).]. Diagnostica. 1997;43(4):347–69.

45. Raine A. The SPQ: A Scale for the Assessment of Schizotypal Personality Based on DSM-III-R Criteria. Schizophr Bull. 1991 Jan 1;17(4):555–64.

46. Baron-Cohen S, Wheelwright S, Skinner R, Martin J, Clubley E. The Autism-Spectrum Quotient (AQ): Evidence from Asperger Syndrome/High-Functioning Autism, Malesand Females, Scientists and Mathematicians. J Autism Dev Disord. 2001 Feb 1;31(1):5–17.

47. Barkhuijsen H, de Beer R, van Ormondt D. Improved algorithm for noniterative time-domain model fitting to exponentially damped magnetic resonance signals. J Magn Reson 1969. 1987 Jul 1;73(3):553–7.

48. Rowland LM, Kontson K, West J, Edden RA, Zhu H, Wijtenburg SA, et al. In Vivo Measurements of Glutamate, GABA, and NAAG in Schizophrenia. Schizophr Bull. 2013 Sep 1;39(5):1096–104.

49. Provencher S. LCModel1 & LCMgui User’s Manual. 2021 Feb 4; Available from: http://lcmodel.ca/pub/LCModel/manual/manual.pdf

50. Maddock RJ, Caton MD, Ragland JD. Estimating Glutamate and Glx from GABA- Optimized MEGA-PRESS: Off-Resonance but not Difference Spectra Values Correspond to PRESS Values. Psychiatry Res Neuroimaging. 2018 Sep 30;279:22–30.

51. Wilson M. Adaptive baseline fitting for MR spectroscopy analysis. Magn Reson Med. 2021;85(1):13–29.

52. Ashburner J, Barnes G, Chen CC, Daunizeau J, Flandin G, Friston K, et al. SPM12 Manual. 2021 Oct 15;

53. Gasparovic C, Song T, Devier D, Bockholt HJ, Caprihan A, Mullins PG, et al. Use of tissue water as a concentration reference for proton spectroscopic imaging. Magn Reson Med. 2006 Jun;55(6):1219–26.

54. Provencher SW. Automatic quantitation of localized in vivo 1H spectra with LCModel. NMR Biomed. 2001;14(4):260–4.

55. Juchem C, Cudalbu C, de Graaf RA, Gruetter R, Henning A, Hetherington HP, et al. B0 shimming for in vivo magnetic resonance spectroscopy: Experts’ consensus recommendations. NMR Biomed. 2021;34(5):e4350.

56. Wilson M, Andronesi O, Barker PB, Bartha R, Bizzi A, Bolan PJ, et al. Methodological consensus on clinical proton MRS of the brain: Review and recommendations. Magn Reson Med. 2019 Aug;82(2):527–50.

57. R Core Team. R: A language and environment for statistical computing [Internet]. Vienna, Austria; 2022. Available from: https://www.R-project.org/

58. Shapiro SS, Wilk MB. An Analysis of Variance Test for Normality (Complete Samples). Biometrika. 1965;52(3/4):591–611.

59. Levene H. Robust tests for equality of variances. Contrib Probab Stat. 1960;278–92.

60. Fox J, Weisberg S. An R companion to applied regression [Internet]. 3rd ed. Thousand Oaks CA: Sage; 2019. Available from: https://socialsciences.mcmaster.ca/jfox/Books/Companion/

61. Friedman M. The Use of Ranks to Avoid the Assumption of Normality Implicit in the Analysis of Variance. J Am Stat Assoc. 1937 Dec 1;32(200):675–701.

62. Kassambara A. rstatix: Pipe-friendly framework for basic statistical tests [Internet]. 2022. Available from: https://CRAN.R-project.org/package=rstatix

63. Rey D, Neuhäuser M. Wilcoxon-Signed-Rank Test. In: Lovric M, editor. International Encyclopedia of Statistical Science [Internet]. Berlin, Heidelberg: Springer; 2011 [cited 2023 Feb 28]. p. 1658–9. Available from: 10.1007/978-3-642-04898-2_616

64. Kassambara A. ggpubr: ‘ggplot2’ based publication ready plots [Internet]. 2022. Available from: https://CRAN.R-project.org/package=ggpubr

65. Kassambara A. ggcorrplot: Visualization of a correlation matrix using ‘ggplot2’ [Internet]. 2022. Available from: https://CRAN.R-project.org/package=ggcorrplot

66. Wuthrich V, Bates TC. Confirmatory Factor Analysis of the Three-Factor Structure of the Schizotypal Personality Questionnaire and Chapman Schizotypy Scales. J Pers Assess. 2006 Oct 1;87(3):292–304.

67. Akoglu H. User’s guide to correlation coefficients. Turk J Emerg Med. 2018 Sep;18(3):91–3.

68. Minati L, Aquino D, Bruzzone M, Erbetta A. Quantitation of normal metabolite concentrations in six brain regions by in-vivo ^1^ H-MR spectroscopy. J Med Phys. 2010;35(3):154.

69. Govindaraju V, Young K, Maudsley AA. Proton NMR chemical shifts and coupling constants for brain metabolites. NMR Biomed. 2000 May;13(3):129–53.

70. Zahid U, Onwordi EC, Hedges EP, Wall MB, Modinos G, Murray RM, et al. Neurofunctional correlates of glutamate and GABA imbalance in psychosis: A systematic review. Neurosci Biobehav Rev. 2023 Jan 1;144:105010.

71. Egerton A, Grace AA, Stone J, Bossong MG, Sand M, McGuire P. Glutamate in schizophrenia: Neurodevelopmental perspectives and drug development. Schizophr Res. 2020 Sep;223:59–70.

72. Near J, Harris AD, Juchem C, Kreis R, Marjańska M, Öz G, et al. Preprocessing, analysis and quantification in single-voxel magnetic resonance spectroscopy: experts’ consensus recommendations. NMR Biomed. 2021 May;34(5):e4257.

73. Dempster K, Jeon P, MacKinley M, Williamson P, Théberge J, Palaniyappan L. Early treatment response in first episode psychosis: a 7-T magnetic resonance spectroscopic study of glutathione and glutamate. Mol Psychiatry. 2020 Aug;25(8):1640–50.

74. Henry ME, Lauriat TL, Shanahan M, Renshaw PF, Jensen JE. Accuracy and stability of measuring GABA, glutamate, and glutamine by proton magnetic resonance spectroscopy: A phantom study at 4Tesla. J Magn Reson. 2011 Feb;208(2):210–8.

