## Supplementary materials for "The impact of spectral basis set composition on estimated levels of cingulate glutamate and its associations with different personality traits"

#### 1 Introduction:

Table S1:

| Metabolites | A | B | C | D | E | F | G | H | I |
|---|---|---|---|---|---|---|---|---|---|
| Ala | | | X | X | X | X | X | X | X |
| Asc | | | X | | X | | | | |
| Asp | X | X | X | X | X | X | X | X | X |
| Ch | X | | | | | | | | |
| Cr | X | X | X | X | X | X | X | X | X |
| GABA | X | X | X | X | X | X | X | X | X |
| Glc | | X | X | X | X | X | X | X | X |
| Gln | X | X | X | X | X | X | X | X | X |
| Glu | X | X | X | X | X | X | X | X | X |
| Gly | | | | | X | X | | X | |
| GSH | | X | X | X | X | X | | X | X |
| GPC | | X | X | X | X | X | X | X | X |
| Lac | | | X | X | X | X | X | X | X |
| mI | X | X | X | X | X | X | X | X | X |
| NAA | X | X | X | X | X | X | X | X | X |
| NAAG | X | X | X | X | X | X | X | X | X |
| PCh | | X | X | | X | X | X | X | X |
| PCr | X | X | X | X | | X | | X | |
| PE | | | X | | X | X | | X | |
| sI | X | X | X | X | X | X | X | X | X |
| Tau | X | X | X | X | X | X | X | X | X |
| GA | | | | | | | X | | |

*Note: Composition of basis sets for the study of A) Cheng et al. (1), B) Maddock et al. (2), C) Reid et al. (3), D) LCModel Manual (4), E) Kozhuharova et al. 2021(5), F) Leptourgos et al. 2023 (6), G) Ford et al. 2017 (7), H) Shukla et al. 2019 (8) and I) Rowland et al. 2013 (9); Ala, alanine; Asc, ascorbate; Asp, aspartate; Cr, creatine; Glc, glucose; Glu, glutamate; Gln, glutamine; Gly, glycine; GSH, glutathione; GPC, glycerophosphocholine; Lac, lactate; mI, myo-Inositol; NAA, N-acetylaspartate; NAAG, N-acetylaspartylglutamate; PCh, phosphocholine; PCr, phosphocreatine; PE, phosphoroylethanolamine; sI, scyllo-Inositol; Tau, taurine; GA, guanidinoacetate. Grey background indicates example basis sets chosen for analysis in this study.*

### 2 Methods

#### 2.1 Participants

*Inclusion criteria:* native German speaker; right-handed; no diagnosis of schizophrenia, psychosis or autism, or any neurological disease or injury; not currently taking any psychoactive medication within at least the past six weeks and no contraindications for MRI scanning.

*Subjects:* Out of the participants, three had been previously diagnosed with depression, two had prior eating disorders, one was diagnosed with schizoid personality disorder along with adjustment disorder, and another reported having post-traumatic stress disorder and narcissistic personality disorder. However, only one of the participants was consistently taking antipsychotic medication (fluoxetine, atomoxetine) for attention deficit disorder.

Details of demographic data and symptom scores are shown in Table S2 and have been previously published in (10).

*Table S2: Demographic data and clinical scores.*

| | Female (n = 26) | Male (n = 27) | P-value[a] | W |
|---|---|---|---|---|
| Age | 23.31 (3.54) | 23.93 (4.21) | 0.7266 | 331 |
| SPQ: Positive-like Symptoms (/132) | 35.96 (23.54) | 28.70 (25.20) | 0.1846 | 426 |
| SPQ: Negative-like Symptoms (/100) | 37.85 (15.57) | 34.33 (20.49) | 0.3454 | 404 |
| SPQ: Disorganized Traits (/64) | 23.19 (14.27) | 21.22 (13.12) | 0.7285 | 371 |
| AQ: Total Score (/50) | 21.65 (6.99) | 20.96 (7.65) | 0.7017 | 373 |

*Note: Values are mean (SD)*

*SPQ, Schizotypal Personality Questionnaire; AQ, Autism Spectrum Quotient*

[a] Wilcoxon rank sum test

#### 2.2 $^{1}$H-MRS Processing

*Figure S1:*

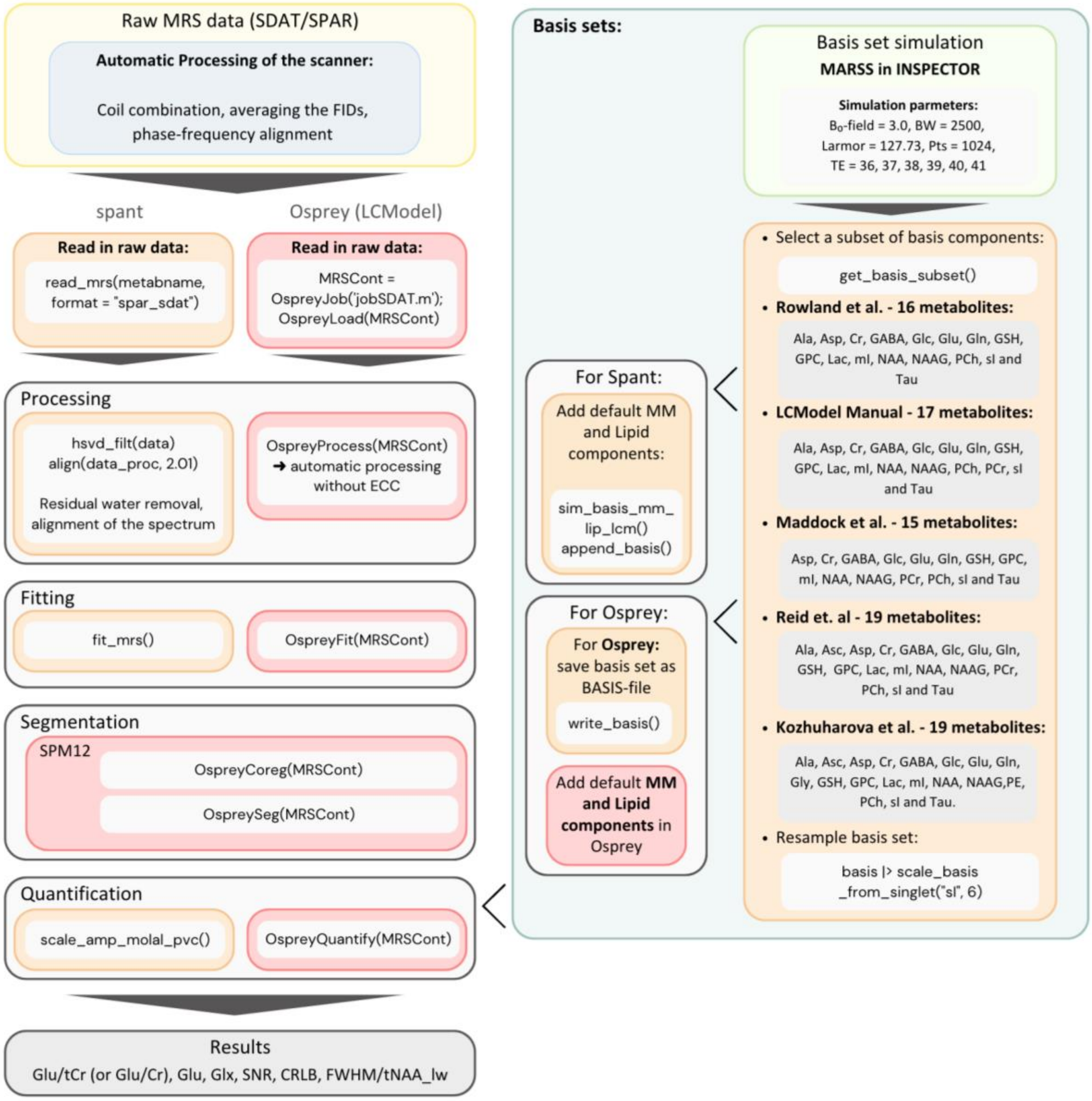


*Note: Processing, modeling, and basisset workflow, summarizing the differences between the applied algorithms and basis sets; colors indicate the toolboxes used: orange, Spant; red, Osprey; green, MARSS in INSPECTOR; MRS, Magnetic resonance spectroscopy; ECC, Eddy current correction; MM, Macromolecules; Ala, alanine; Asc, ascorbate; Asp, aspartate; Cr, creatine; Glc, glucose; Glu, glutamate; Gln, glutamine; Gly, glycine; GSH, glutathione; GPC, glycerophosphocholine; Lac, lactate; ml, myo-Inositol; NAA, N-acetylaspartate; NAAG, N-acetylaspartylglutamate; PCh, phosphocholine; PCr, phosphocreatine; PE, phosphoroylethanolamine; sl, scyllo-Inositol; Tau, taurine; SNR, Signal-to-Noise Ratio; CRLB, Cramer-Rao lower bounds; FWHM, full width at half maximum, measurement for the water linewidth; tNAA_lw, measurement for the tNAA (NAA + NAAG) linewidth*

#### 3 Results

##### 3.1 Spectral quality

We applied the paired Wilcoxon signed-rank test for multiple pairwise comparisons of the CRLB and the multiple pairwise paired t-tests for the SNR between basis sets. P-values were adjusted using the Bonferroni multiple-testing correction method. An adjusted p-value of less than 0.05 was considered significant. Results are presented in Table S3 and Table S4.

*Table S3: Summary spectral quality parameter distributions of the basis sets*

| Quality Parameter | | Rowland | LCModel | Maddock | Reid | Kozhuha. | Group comparisons | |
|---|---|---|---|---|---|---|---|---|
| Analysis method | n | Mean ± SD | Mean ± SD | Mean ± SD | Mean ± SD | Mean ± SD | Statistic | P-value |
| **Glu CRLB (%)** | | | | | | | | |
| Spant + ABfit | 53 | 4.04 ± 0.88 | 3.79 ± 0.74 | 3.87 ± 0.68 | 3.62± 0.86 | 3.83 ± 0.78 | $X^2_4$ = 40.33[a] | <0.0001 |
| Osprey + LCM | 53 | 5.32 ± 0.58 | 5.25 ± 0.52 | 5.25 ± 0.52 | 5.09 ± 0.66 | 5.25 ± 0.55 | $X^2_4$ = 17.45[a] | 0.0016 |
| **Glx CRLB (%)** | | | | | | | | |
| Spant + ABfit | 53 | 1.83 ± 0.67 | 1.77 ± 0.51 | 1.89 ± 0.47 | 1.81 ± 0.59 | 1.62 ± 0.56 | $X^2_4$ = 12.40[a] | 0.0146 |
| Osprey + LCM | 53 | 5.17 ± 0.67 | 5.21 ± 0.50 | 5.23 ± 0.51 | 4.98 ± 0.69 | 4.93 ± 0.76 | $X^2_4$ = 27.47[a] | <0.0001 |
| **SNR** | | | | | | | | |
| Spant + ABfit | 53 | 149.18 ± 18.01 | 148.02 ± 18.33 | 147.88 ± 18.20 | 149.23 ± 18.36 | 149.89 ± 18.05 | $F_{1.68, 87.49}$ = 31.03[b] | <0.0001 |

| | | | | | | | | |
|---|---|---|---|---|---|---|---|---|
| Osprey + LCM | 53 | 101.06 ± 13.88 | 101.06 ± 13.88 | 101.06 ± 13.88 | 101.06 ± 13.88 | 101.06 ± 13.88 | NaN[b] | NaN |
| **FWHM (ppm)** | | | | | | | | |
| Spant + ABfit | 53 | 0.042 ± 0.01 | 0.041 ± 0.01 | 0.041 ± 0.01 | 0.041 ± 0.01 | 0.042 ± 0.01 | $X^2_4$ = 150.7[a] | **<0.0001** |
| **FWHM (Hz)** | | | | | | | | |
| Osprey + LCM | 53 | 6.18 ± 0.44 | 6.18 ± 0.44 | 6.18 ± 0.44 | 6.18 ± 0.44 | 6.18 ± 0.44 | NaN [a] | NaN |

*Note: Kozhuha.., Kozhuharova; SNR, Signal-to-Noise Ratio; CRLB, Cramer-Rao lower bounds; FWHM, full width at half maximum, in Spant: measurement for the tNAA (NAA + NAAG) linewidth in Osprey: measurement for the water linewidth*

[a] Calculated using the Friedman test

[b] Calculated using the repeated measures ANOVA

*Table S4: Pairwise comparisons for the SNR*

| | | *spant+ABfit* | | *Osprey+LCM* | |
|---|---|---|---|---|---|
| **group1** | **group2** | **statistic** | **P-value adj.** | **statistic** | **P-value adj.** |
| Rowland | LCModel | 5.20 | **<0.0001** | NA | NA |
| Rowland | Maddock | 5.57 | **<0.0001** | NA | NA |
| Rowland | Reid | -0.24 | 1 | NA | NA |
| Rowland | Kozhuharova | -5.03 | **<0.0001** | NA | NA |
| LCModel | Maddock | 1.88 | 0.662 | NA | NA |
| LCModel | Reid | -8.84 | **<0.0001** | NA | NA |
| LCModel | Kozhuharova | -6.76 | **<0.0001** | NA | NA |
| Maddock | Reid | -9.36 | **<0.0001** | NA | NA |
| Maddock | Kozhuharova | -6.84 | **<0.0001** | NA | NA |

| | | | | | |
|---|---|---|---|---|---|
| Reid | Kozhuharova | -2.27 | 0.272 | NA | NA |

Note: Multiple pairwise paired t-tests for the SNR of spant+ABfit and Osprey+LCM analyses; SNR, Signal-to-Noise Ratio; P-value adj.; adjusted p-value using the Bonferroni multiple-testing correction.

*Table S5: Pairwise comparisons for Glu CRLB*

| | | *spant+ABfit* | | *Osprey+LCM* | |
|---|---|---|---|---|---|
| **group1** | **group2** | **statistic** | **P-value adj.** | **statistic** | **P-value adj.** |
| Rowland | LCModel | 352 | 0.201 | 38.5 | 1 |
| Rowland | Maddock | 285 | 0.971 | 38.5 | 1 |
| Rowland | Reid | 409 | **0.005** | 97.5 | **0.014** |
| Rowland | Kozhuharova | 85.5 | 0.277 | 17.5 | 1 |
| LCModel | Maddock | 3.5 | 1 | 0 | NA |
| LCModel | Reid | 66 | 0.226 | 49.5 | 0.121 |
| LCModel | Kozhuharova | 138.5 | 1 | 18 | 1 |
| Maddock | Reid | 85 | **0.034** | 49.5 | 0.121 |
| Maddock | Kozhuharova | 188 | 1 | 18 | 1 |
| Reid | Kozhuharova | 94 | 0.459 | 13 | 0.212 |

Note: Wilcoxon signed-rank test pairwise comparisons for the glutamate CRLB of spant+ABfit and Osprey+LCM analyses; CRLB, Cramer-Rao lower bounds; P-value adj., adjusted p-value using the Bonferroni multiple-testing correction.

*Table S6: Pairwise comparisons for Glx CRLB*

| | | *spant+ABfit* | | *Osprey+LCM* | |
|---|---|---|---|---|---|
| **group1** | **group2** | **statistic** | **P-value adj.** | **statistic** | **P-value adj.** |
| Rowland | LCModel | 132 | 1 | 45 | 1 |

| | | | | | |
|---|---|---|---|---|---|
| Rowland | Maddock | 120 | 1 | 35 | 1 |
| Rowland | Reid | 121 | 1 | 110.5 | 0.135 |
| Rowland | Kozhuharova | 79 | 0.132 | 98 | **0.018** |
| LCModel | Maddock | 4.5 | 0.411 | 0 | 1 |
| LCModel | Reid | 13.5 | 1 | 119 | **0.03** |
| LCModel | Kozhuharova | 123.5 | 0.628 | 228 | **0.019** |
| Maddock | Reid | 38.5 | 1 | 135 | **0.018** |
| Maddock | Kozhuharova | 178.5 | **0.019** | 218.5 | **0.007** |
| Reid | Kozhuharova | 142.5 | 0.349 | 110 | 1 |

Note: Wilcoxon signed-rank test pairwise comparisons for the Glx CRLB of spant+ABfit and Osprey+LCM analyses; CRLB, Cramer-Rao lower bounds; P-value adj., adjusted p-value using the Bonferroni multiple-testing correction.

*Table S7: FWHM*

| | | *spant+ABfit* | | *Osprey+LCM* | |
|---|---|---|---|---|---|
| **group1** | **group2** | **statistic** | **P-value adj.** | **statistic** | **P-value adj.** |
| Rowland | LCModel | 1428 | **<0.0001** | NA | NA |
| Rowland | Maddock | 1428 | **<0.0001** | NA | NA |
| Rowland | Reid | 1430 | **<0.0001** | NA | NA |
| Rowland | Kozhuharova | 966 | 0.269 | NA | NA |
| LCModel | Maddock | 723 | 1 | NA | NA |
| LCModel | Reid | 994 | 0.139 | NA | NA |
| LCModel | Kozhuharova | 4 | **<0.0001** | NA | NA |
| Maddock | Reid | 819 | 1 | NA | NA |
| Maddock | Kozhuharova | 1 | **<0.0001** | NA | NA |

| | | | | | |
|---|---|---|---|---|---|
| Reid | Kozhuharova | 0 | <0.0001 | NA | NA |

Note: Wilcoxon signed-rank test pairwise comparisons for the FWHM of tNAA for spant+ABfit and of water for Osprey+LCM analyses; FWHM, full width at half maximum; P-value adj., adjusted p-value using the Bonferroni multiple-testing correction.

##### 3.2 Group differences between the metabolite concentration estimates

Comparisons between the estimates for Glu/tCr, Glu, and Glx are summarized in **Figure 5**, results are presented in **Table S8-S11.**

*Table S8: Differences between the metabolite concentration distributions of the basis sets*

| Metabolite | | Rowland | LCModel | Maddock | Reid | Kozhuha. | Group comparisons | |
|---|---|---|---|---|---|---|---|---|
| Analysis method | n | Mean ± SD | Mean ± SD | Mean ± SD | Mean ± SD | Mean ± SD | Statistic | P-value |
| **Glu/tCR** | | | | | | | | |
| Spant + ABfit | 53 | 1.71 ± 0.22 | 1.49 ± 0.22 | 1.46 ± 0.22 | 1.46 ± 0.27 | 1.76 ± 0.22 | $X^2_4$ = 72.15 | <0.0001 |
| Osprey + LCM | 53 | 1.69 ± 0.11 | 1.56 ± 0.10 | 1.55 ± 0.10 | 1.53 ± 0.11 | 1.68 ± 0.10 | $X^2_4$ = 166.88 | <0.0001 |
| **Glu** | | | | | | | | |
| Spant + ABfit | 53 | 20.53 ± 2.87 | 18.18 ± 2.95 | 17.72 ± 2.93 | 18.01 ± 3.54 | 21.48 ± 2.86 | $X^2_4$ = 79.98 | <0.0001 |
| Osprey + LCM | 53 | 30.21 ± 1.93 | 28.12 ± 1.91 | 28.01 ± 1.90 | 28.13 ± 1.89 | 30.13 ± 1.81 | $X^2_4$ = 151.02 | <0.0001 |
| **Glx** | | | | | | | | |
| Spant + ABfit | 53 | 24.28 ± 3.34 | 21.28 ± 3.23 | 20.81 ± 3.21 | 21.37 ± 4.01 | 26.39 ± 4.01 | $X^2_4$ = 89.51 | <0.0001 |

| | | | | | | | | |
|---|---|---|---|---|---|---|---|---|
| Osprey + LCM | 53 | 36.56 ± 2.31 | 33.37 ± 2.07 | 33.28 ± 2.06 | 33.20 ± 216 | 36.50 ± 2.17 | $X^2_4$ = 149.69 | <0.0001 |

Note: Statistics are calculated using the Friedman test; all values were rounded to two decimal places. For Glu/tCr the glutamate concentration is referenced to tCr, except for Rowland and Kozhuharova (Glu/Cr). The absolute Glu and Glx values are given in mol/kg. Kozhuha., Kozhuharova; Glu, glutamate; tCr, total creatin (creatin + phosphocreatine); Glx, glutamate + glutamine

*Table S9: Pairwise comparisons for Glu/tCr*

| | | spant+ABfit | | Osprey+LCM | |
|---|---|---|---|---|---|
| **group1** | **group2** | **statistic** | **P-value adj.** | **statistic** | **P-value adj.** |
| Rowland | LCModel | 1309 | **<0.0001** | 1428 | **<0.0001** |
| Rowland | Maddock | 1329 | **<0.0001** | 1429 | **<0.0001** |
| Rowland | Reid | 1292 | **<0.0001** | 1428 | **<0.0001** |
| Rowland | Kozhuharova | 491 | 0.474 | 1224 | **<0.0001** |
| LCModel | Maddock | 936 | 0.515 | 1406 | **<0.0001** |
| LCModel | Reid | 977 | 0.209 | 1256 | **<0.0001** |
| LCModel | Kozhuharova | 53 | **<0.0001** | 10 | **<0.0001** |
| Maddock | Reid | 777 | 1 | 1172 | **0.0005** |
| Maddock | Kozhuharova | 59 | **<0.0001** | 4 | **<0.0001** |
| Reid | Kozhuharova | 98 | **<0.0001** | 8 | **<0.0001** |

Note: Wilcoxon signed-rank test pairwise comparisons for Glu/tCr of spant+ABfit and Osprey+LCM analyses; Glu, glutamate; tCr, total creatin (creatin + phosphocreatine); P-value adj.; adjusted p-value using the Bonferroni multiple-testing correction.

*Table S10: Pairwise comparisons for Glu*

| | | spant+Abfit | | Osprey+LCM | |
|---|---|---|---|---|---|
| **group1** | **group2** | **statistic** | **P-value adj.** | **statistic** | **P-value adj.** |
| Rowland | LCModel | 1268 | <0.0001 | 1426 | <0.0001 |
| Rowland | Maddock | 1303 | <0.0001 | 1429 | <0.0001 |
| Rowland | Reid | 1186 | 0.0003 | 1420 | <0.0001 |
| Rowland | Kozhuharova | 277 | 0.001 | 936 | 0.515 |
| LCModel | Maddock | 1055 | 0.027 | 1370 | <0.0001 |
| LCModel | Reid | 873 | 1 | 719 | 1 |
| LCModel | Kozhuharova | 32 | <0.0001 | 5 | <0.0001 |
| Maddock | Reid | 598 | 1 | 551 | 1 |
| Maddock | Kozhuharova | 34 | <0.0001 | 2 | <0.0001 |
| Reid | Kozhuharova | 128 | <0.0001 | 19 | <0.0001 |

Note: Wilcoxon signed-rank test pairwise comparisons for absolute Glu values of spant+ABfit and Osprey+LCM analyses; Glu, glutamate; P-value adj.; adjusted p-value using the Bonferroni multiple-testing correction.

*Table S11: Pairwise comparisons for Glx*

| | | spant+Abfit | | Osprey+LCM | |
|---|---|---|---|---|---|
| **group1** | **group2** | **statistic** | **P-value adj.** | **statistic** | **P-value adj.** |
| Rowland | LCModel | 1295 | <0.0001 | 1429 | <0.0001 |

| | | | | | |
|---|---|---|---|---|---|
| Rowland | Maddock | 1324 | <0.0001 | 1429 | <0.0001 |
| Rowland | Reid | 1166 | 0.0006 | 1424 | <0.0001 |
| Rowland | Kozhuharova | 118 | <0.0001 | 802 | 1 |
| LCModel | Maddock | 1030 | 0.054 | 1232 | <0.0001 |
| LCModel | Reid | 748 | 1 | 940 | 0.474 |
| LCModel | Kozhuharova | 9 | <0.0001 | 1 | <0.0001 |
| Maddock | Reid | 536 | 1 | 855 | 1 |
| Maddock | Kozhuharova | 8 | <0.0001 | 0 | <0.0001 |
| Reid | Kozhuharova | 118 | <0.0001 | 5 | <0.0001 |

Note: Wilcoxon signed-rank test pairwise comparisons for absolute Glx values of spant+ABfit and Osprey+LCM analyses; Glx, glutamate+glutamine; P-value adj.; adjusted p-value using the Bonferroni multiple-testing correction.

### 4 Discussion

#### 4.1 Group differences between the metabolite concentration estimates

Comparisons between the estimates for Glx are summarized in **Figure S2**. Adding PCr to the basis sets leads to a decrease in the Glx concentrations for both the Rowland(9) and the Kozhuharova(5) basis sets as well as in both toolboxes. Estimates were consistently lower for the analysis in spant+ABfit compared to the ones analyzed with Osprey+LCM. Necessary to note is also that the Rowland+PCr and LCModel basis sets are now identical, which can be nicely seen in the boxplots.

#### 4.2 Correlations of each metabolite estimate between the basis sets

As already mentioned in the manuscript, the correlations between metabolite (Glu/tCr, absolute Glu, Glx) and basis set per toolbox revealed that the metabolite concentrations between the basis sets had higher correlations using Ospreys LCM integration than spant+ABfit. For Osprey+LCM, we found strong correlations between all basis sets (r>0.75). Comparing the Rowland (9) and Kozhuharova (5) basis sets with added PCr to the ones without, the “new” basis sets show higher correlations with the three visually best-fitting basis sets (LCModel Manual (4), Maddock et al. (2), and Reid et al. (3); **Figure 3 B, C, and D**) with Spearman correlation coefficients between 0.95-1 for Osprey+LCM and 0.87-0.97 for spant+ABfit, the three visually best-fitting basis sets (LCModel Manual (4), Maddock et al. (2), and Reid et al. (3); **Figure 3 B, C, and D**) showed the best results with spearman correlation coefficients between 0.93-1 for Osprey+LCM and 0.88-0.94 for spant+ABfit (**Figure S3**). Correlation strength was classified according to Akoglu (11).

##### 4.3 Association between the concentrations and traits

Finally, we also analyzed Spearman's rank correlations for clinical scores with the extracted concentration scores for the two basis sets with added PCr. Overall, the results in Osprey+LCM and ant+ABfit displayed now a more homogenous pattern regarding their correlations. Using the Rowland (9)and Kozhuharova(5) basis set with PCr instead of the ones without this metabolite, the maximum difference between the correlation coefficients of Glx and disorganized traits in Osprey+LCM decreased from 0.18 to 0.04. The same can be seen in spant+ABfit where the maximal difference was changing from 0.42 to 0.16 for this correlation. However, the difference between the toolboxes was still smaller than the difference across the different basis sets and reached a maximum of 0.11, again for the correlation of Glx and disorganized traits. The correlations are shown in **Figure S2.**

*Figure S2: Basisets + PCr: Group comparisons for Glx in the ACC and Correlations between the metabolite concentrations and clinical scores*

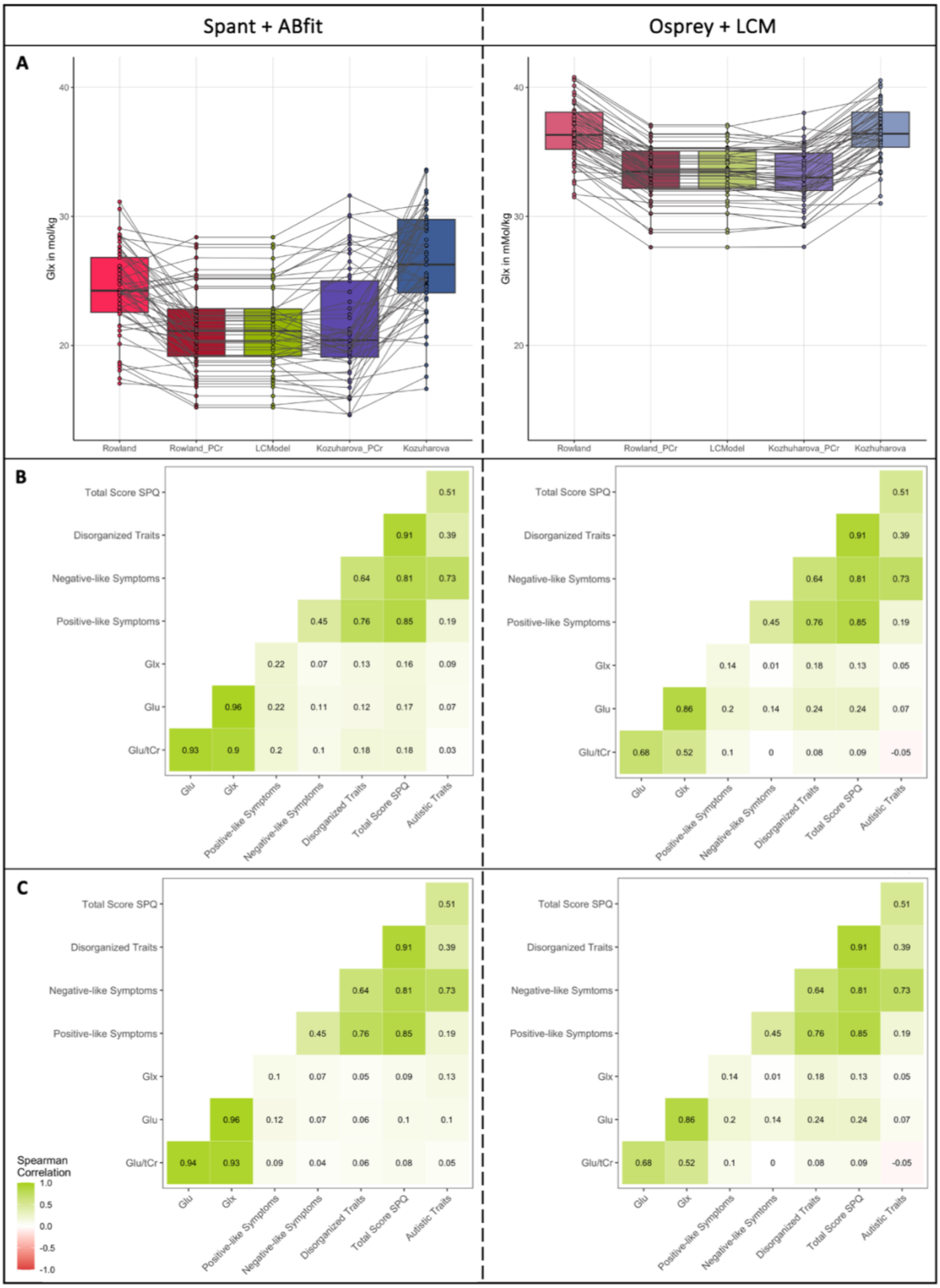


*Note: A) Group comparisons between the basis sets (Rowland* (12)*, Rowland + PCr, LCModel* (4)*, Kozhuharova +PCr, Kozhuharova* (5) *for absolute Glx values; Spearman correlations between Glu/tCr (Glu/Cr for Rowland and Kozhuharova), absolute Glu, Glx, and the subclinical traits for the basis sets B)*

*Rowland + PCr and C)* (5)*; the left side always shows the results for spant+ABfit and the right side for Osprey+LCM. The psychotic-like traits are separated into the subscores: positive-like symptoms, negative-like symptoms, and disorganized traits.*

*Figure S3: Basis sets + PCr and tool box intercorrelations*

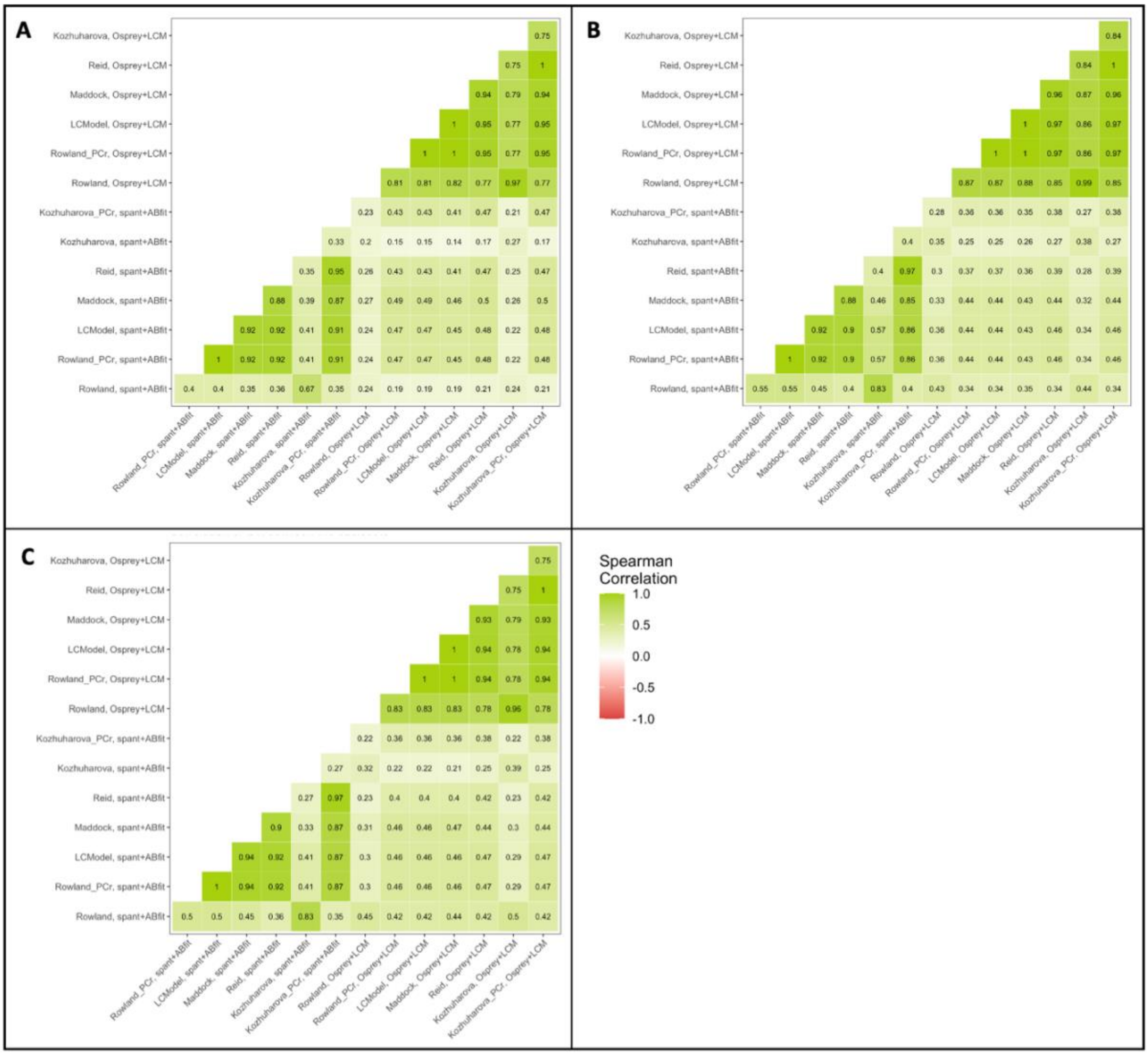


*Note: Intercorrelations between the basis sets (Rowland* (12)*, Rowland+PCr, LCModel* (4)*, Maddock* (2)*, Reid* (3)*, Kozhuharova* (5) *and Kozhuharova+PCr) and toolboxes (spant+ABfit; Osprey) for A) Glu/tCr (Glu/Cr for Rowland and Kozhuharova), B) Glu, C) Glx*

References:


1. *Cheng H, Wang A, Newman S, Dydak U. An investigation of glutamate quantification with PRESS and MEGA-PRESS. NMR in Biomedicine. 2021;34(2):e4453.*

2. *Maddock RJ, Caton MD, Ragland JD. Estimating Glutamate and Glx from GABA-Optimized MEGA-PRESS: Off-Resonance but not Difference Spectra Values Correspond to PRESS Values. Psychiatry Res Neuroimaging. 2018 Sep 30;279:22–30.*

3. *Reid MA, Salibi N, White DM, Gawne TJ, Denney TS, Lahti AC. 7T Proton Magnetic Resonance Spectroscopy of the Anterior Cingulate Cortex in First-Episode Schizophrenia. Schizophr Bull. 2019 Jan 1;45(1):180–9.*

4. *Provencher S. LCModel1 & LCMgui User's Manual. 2021 Feb 4; Available from: http://lcmodel.ca/pub/LCModel/manual/manual.pdf*

5. *Kozhuharova P, Diaconescu AO, Allen P. Reduced cortical GABA and glutamate in high schizotypy. Psychopharmacology. 2021 Sep;238(9):2459–70.*

6. *Leptourgos P, Bansal S, Dutterer J, Culbreth A, Powers A III, Suthaharan P, et al. Relating Glutamate, Conditioned, and Clinical Hallucinations via 1H-MR Spectroscopy. Schizophrenia Bulletin. 2022 Jul 1;48(4):912–20.*

7. *Ford TC, Nibbs R, Crewther DP. Glutamate/GABA+ ratio is associated with the psychosocial domain of autistic and schizotypal traits. PLOS ONE. 2017 Jul 31;12(7):e0181961.*

8. *Shukla DK, Wijtenburg SA, Chen H, Chiappelli JJ, Kochunov P, Hong LE, et al. Anterior Cingulate Glutamate and GABA Associations on Functional Connectivity in Schizophrenia. Schizophrenia Bulletin. 2019 Apr 25;45(3):647–58.*

9. *Rowland LM, Kontson K, West J, Edden RA, Zhu H, Wijtenburg SA, et al. In Vivo Measurements of Glutamate, GABA, and NAAG in Schizophrenia. Schizophrenia Bulletin. 2013 Sep 1;39(5):1096–104.*

10. *Demler VF, Sterner EF, Wilson M, Zimmer C, Knolle F. Association between increased anterior cingulate glutamate and psychotic-like symptoms, but not autistic traits [Internet]. medRxiv; 2023 [cited 2023 Mar 6]. p. 2023.02.01.23285183. Available from: https://www.medrxiv.org/content/10.1101/2023.02.01.23285183v1*

11. *Akoglu H. User's guide to correlation coefficients. Turkish Journal of Emergency Medicine. 2018 Sep;18(3):91–3.*

12. *Rowland LM, Kontson K, West J, Edden RA, Zhu H, Wijtenburg SA, et al. In Vivo Measurements of Glutamate, GABA, and NAAG in Schizophrenia. Schizophrenia Bulletin. 2013 Sep 1;39(5):1096–104.*
